# The Growth Hormone-IGF-1 axis is a risk factor for Long-Term Kidney Allograft Failure

**DOI:** 10.1101/2024.11.27.24318002

**Authors:** Matthew Cusick, Viji Nair, Damian Fermin, John Hartman, Jeffrey A. Beamish, Zeguo Sun, Zhongyang Zhang, Edgar Otto, Rajasree Menon, Sudha Nadimidla, Nicholas Demchuk, Kelly Shaffer, Peter Heeger, Weija Zhang, Madhav C. Menon, Matthias Kretzler, Roger C. Wiggins, Abhijit S. Naik

## Abstract

**Introduction:** Maladaptive hypertrophy and podocyte stress and depletion contribute to kidney function decline. Although IGF-1 plays a key role in early hypertrophic responses in the single kidney state, its impact on KTx outcomes remains uncertain. This report tests the hypothesis that early IGF-1 exposure reduces KTx survival.

**Methods:** Population datasets compared incident Death Censored Graft Failure (DCGF) rates by age at KTx (n=366,404) with IGF-1 levels by age (n=15,014). A clinical study of 216 KTx recipients evaluated the association of IGF-1 exposure with DCGF and secondary outcomes of proteinuria and Biopsy-Proven Acute Rejection. IGF-1 exposure was modeled using pre-KTx IGF-1 levels and donor kidney dose estimated from the donor:recipient body surface area ratio reflecting allograft hyperfiltration. The association of DCGF with an IGF1 SNP linked to high IGF-1 levels was assessed in 724 genotyped allograft recipients. Single-cell transcriptomic data from first-year post-KTx patients (n=14) and binephric donors (n=18) were compared to assess intrarenal cellular expression of *IGF1, IGF1R, and GHR* transcripts.

**Results:** DCGF risk by age at KTx paralleled IGF-1 levels by age. Higher IGF-1 exposure was associated with significantly increased risks of DCGF, proteinuria and T-Cell mediated rejection. Genotypic analysis showed a 50% increase in DCGF risk per risk allele at IGF1 eQTL *rs35767*. First-year biopsy results revealed no increase in intrarenal *IGF1* transcript, while *GHR* and *IGF-1R* transcripts were suppressed, consistent with circulating IGF-1 (vs. graft-derived IGF-1) being the primary source of IGF-1 exposure.

**Conclusion:** We identify a novel role for the GH-IGF-1 axis in reducing KTx survival.

## Introduction

Reduced kidney mass scenarios such as 5/6^th^ nephrectomy, which progresses rapidly, and unilateral nephrectomy (Uni-Nx), which progresses more slowly to kidney failure, develop from adaptations to reduced renal mass(1,2). These adaptations include hyperfiltration driven by nitric oxide-mediated vasodilatation(3), podocyte hypertrophic stress, and depletion, culminating in proteinuria, glomerulosclerosis, and End-Stage Kidney Disease (ESKD)(4,5). Thus, adaptative or maladaptive hypertrophic responses to reduced renal mass can influence long-term kidney outcomes(6,7).

All single kidney allografts in KTx recipients are forced to hyperfilter and hypertrophy to meet the metabolic demands of the host due to the lower kidney mass (dose) typically received at Kidney Transplantation (KTx)(8). In addition to a single kidney replacing the normal two-kidney complement, a lower body surface area (BSA) of the donor versus that of the recipient (reflecting a lower kidney dose) is associated with inferior KTx survival(9,10). Our previous reports also highlight a critical role of kidney dose in KTx outcomes, with smaller kidney doses associated with elevated urine markers of podocyte stress, detachment, and accelerated graft function decline aligning with UniNx studies(4,5,11,12). Notably, podocyte density significantly decreased by 20% by 3- and 30% by 12 months post-KTx(13). This level of podocyte density reduction in model systems(14) and human studies(15) is associated with glomerular destabilization, glomerulosclerosis, and reduced long-term kidney survival(11–13,16). Consistent with this hypothesis, we have previously shown that even stable allografts have a podocyte detachment rate that is approximately six-fold higher than binephric healthy controls(13). Thus, an early hypertrophic but potentially maladaptive response to reduced kidney dose is associated with excess podocyte stress and detachment that can induce podocyte depletion, culminating in shortened KTx survival(9,10,17–19). Elucidating the key mediators that play a role in this early hypertrophic response and identifying the factors driving towards a maladaptive response could offer potentially targetable candidate pathways to improve long-term KTx outcomes.

The Growth Hormone (GH) and Insulin-like Growth Factor-1 (IGF-1) axis play a fundamental role in post-natal organ growth and post-nephrectomy kidney hypertrophy(20–22). Data from several studies, including the use of IGF-1R blockers, support the concept that IGF-1 drives the very early kidney growth response to UniNx, which can become maladaptive(23–26). Furthermore, our work in model systems demonstrates that the rate of remaining kidney growth is maximal by two days post-UniNx, coinciding with the peak in urinary IGF-1 excretion and a simultaneous reduction in circulating IGF-1 level(27). We demonstrate that this very early IGF-1-driven growth profoundly accelerates glomerular growth and podocyte density reduction in the remnant kidney which has long-term consequences(27). Other groups have also demonstrated increased IGF-1 protein content in remnant kidneys early after Uni-Nx(28,29), aligning with the observed increase in early increase in urinary IGF-1 excretion reported previously(27). However, whether this early increase in urine IGF1 is related to kidney cell intrinsic IGF1 overproduction is unclear.

In this report, using population-, clinical, and genetic level data, we first test the hypothesis and demonstrate that high level *IGF-1 exposure from the circulation* in the peri-KTx single kidney state is associated with inferior outcomes, comparable to what is observed in partial nephrectomy model systems of kidney failure(1,4,5). Additionally using single-cell RNA-seq studies in a cohort of kidney transplant recipients in the first post-transplant year, we report that IGF-1 transcript is not increased to levels above binephric healthy human controls, and that GHR and IGF-1R transcripts are decreased(30). This last analysis further buttresses the hypothesis that circulating IGF-1, as opposed to increased intragraft expression of IGF-1 is responsible for "compensatory" adaptations in clinically stable allografts and this could offer a potentially targetable pathway to mitigate early injury.

## Results

Figure 1 shows the multicohort approach that was utilized to test the hypothesis. Initially, we made use of the fact that IGF-1 levels change by 6-fold during a normal lifespan to test the hypothesis that allograft survival would be linked to normal changes in IGF-1 level. We utilized a publicly available population dataset to examine the relationship between IGF-1 and age using a reference IGF-1 report(31). In parallel we evaluated long-term death censored graft failure (DCGF) rates by age at KTx using the Organ Procurement and Transplant Network (OPTN) dataset(32). We hypothesized that if circulating (blood) IGF-1 levels were a driver of long-term kidney allograft failure, then we should observe a relationship between IGF-1 levels and long-term kidney allograft failure by age anticipating that at ages of highest IGF-1 levels, the graft failure risk should be the highest and *vice versa*. This population-based study was followed by a more granular clinical study wherein we measured individual IGF-1 levels in patients before KTx and linked them to DCGF rates. Additionally, we reasoned that if high blood IGF-1 levels drive DCGF, then any genetically driven increase in blood IGF-1 levels in the kidney transplant recipient should also be associated with higher DCGF rates. To test this hypothesis, from literature curation, we identified a Single Nucleotide Polymorphism (SNP) in the promoter region of *the IGF1* gene, linked to elevated circulating IGF-1 levels. We then tested whether this variant was associated with increased DCGF rates in KTx receipients(33–35). Lastly, using our recently published transcriptomic dataset of healthy compensated kidney allograft biopsies and binephric living kidney donors(30), we explored critical elements of IGF-1 signaling. The rationale for this last study was to determine whether intrarenal IGF-1 mediated signal transduction pathways or circulating IGF-1 contributed to the higher allograft IGF-1 exposure.

**Figure 1.**
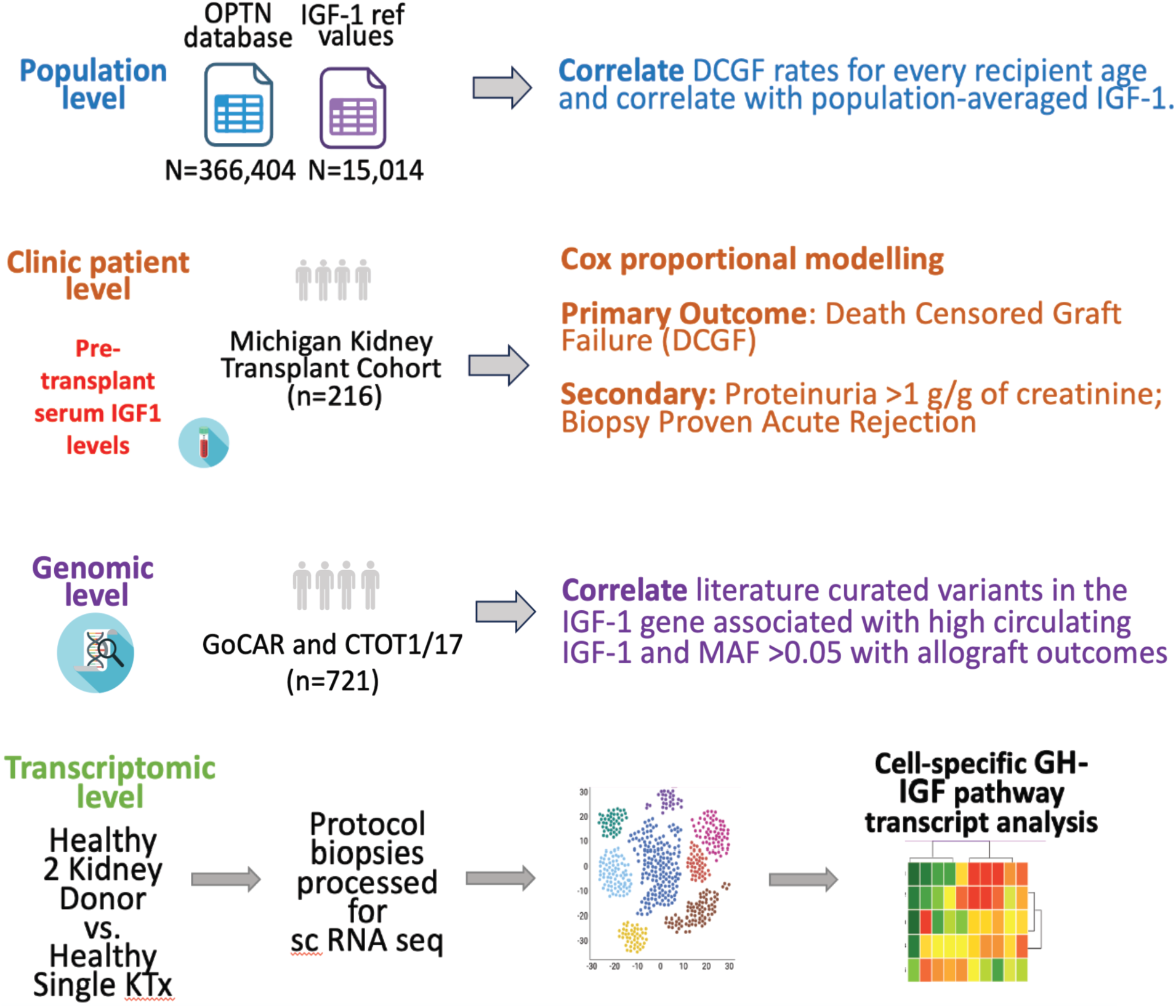
Legend: Approaches used to assess the relationship between IGF-1 levels at kidney transplantation and long-term allograft survival. ***Cohort 1: Population-level study:*** Population-level analysis compared death-censored graft failure rates and time to 50% graft loss among kidney transplant recipients aged 0 −60 with IGF1 levels by age derived from a reference study. ***Cohort 2: Clinical Study*** This analysis involved 216 consecutive kidney transplant recipients aged 0-80 years. Pre-transplant IGF-1 levels were measured and related to long-term outcomes using survival models. These models were adjusted for donor, recipient, and transplant characteristics, including an estimated kidney dose (eKD) derived from the donor-recipient body surface area ratio (see Results section). Adjustments in the Cox survival model included primary outcomes (death-censored graft failure, DCGF), secondary outcomes (Proteinuria>1 g/g), and a composite of alloimmune responses (acute rejection or de novo donor-specific antibody development). ***Cohort 3: Genotype Study:*** For genetic level analysis, two National Institutes of Health-sponsored studies linking genotype to outcomes in kidney transplant recipients were employed to test the hypothesis that an IGF-1 gene variant associated with high circulating IGF-1 levels correlates with long-term allograft survival. **Cohort 4. Transcriptomic study:** Our previously published cohort was utilized for transcriptomic level analysis to map IGF system transcript expression in single kidney cells derived from normal kidney allografts with no histologic abnormality in the first year post-transplant, compared to normal kidneys biopsied at the time of transplantation(30).

### Cohort 1: Population-Level Study

As summarized in Supplemental Figure 1 we used the OPTN dataset comprising 366,404 single kidney transplant recipients including all kidney transplants performed in the USA between 1987 and March 2022. Incident DCGF rates at the age at which the patient underwent KTx were calculated. Additionally, we generated incident DCGF rates based on the time after transplantation by age for short-term (1-year), intermediate-term (3-year), and long-term (10-year) outcomes. We also calculated the time by which 50% of grafts had failed without death (median allograft survival). **Table 1** provides the donor and recipient characteristics of this cohort. Population-based IGF-1 levels by age were obtained from the report of Bidlingmaier et al(31).

**Table 1:** Donor, recipient, and transplant characteristics of the OPTN cohort used for analysis (n=366,404).

|  |  |
| --- | --- |
| **Donor Factors** |  |
| **Donor Age** | 39.1 ±12.6 |
| **Donor Type** |  |
| Living | 141,584 (38.6%) |
| Deceased non-ECD | 170,122 (46.4%) |
| Deceased ECD | 21,973 (6.0%) |
| Missing | 32,725 (8.9%) |
| **Donor Gender** |  |
| Female | 171,171 (46.7%) |
| Male | 195,232 (53.3%) |
| **Donor Race** |  |
| White | 260,011 (71.0%) |
| *African American* | 43,355 (11.8%) |
| Others or missing | 63,038 (17.2%) |
| **Donor BMI** (Missing %=16.7) | 27.2± 5.8 |
| **Donor BSA** (Missing %=16.6) | 1.94± 0.26 |
| **Transplant Factors** |  |
| **Cold Ischemia Factor** |  |
| <12 hours | 158,786 (43.3%) |
| >12 hours | 168,802 (46.1) |
| Missing | 38,816 (10.6%) |
| **HLA Mismatch** |  |
| 0 | 31,214 (8.5%) |
| 1 | 15,993 (4.4%) |
| 2 | 38,366 (10.5%) |
| 3 | 73,301(20.0%) |
| 4 | 78,655 (21.5%) |
| 5 | 83,730 (22.9%) |
| 6 | 41,030 (11.2%) |
| Missing | 4,115 (1.1%) |
| **Recipient Factors** |  |
| **Recipient Age** (years) | 41.0 ± 13.3 |
| **Recipient BMI** (Missing %= 4.7) | 26.20 ± 5.72 |
| **Recipient Gender** |  |
| Female | 145,549 (39.7%) |
| Male | 220,855 (60.3%) |
| **Recipient Race** |  |
| White | 193,721 (52.9%) |
| *African American* | 92,449 (25.2%) |
| Others | 80,234 (21.9%) |
| **Recipient PRA** |  |
| PRA <20 | 245,415 (67.0%) |
| PRA>20 | 67,789 (18.5%) |
| Missing | 81,608 (14.5%) |
| **Induction** |  |
| No | 74,902 (20.4%) |
| Yes | 291,502 (79.6%) |
| **Maintenance** |  |
| CNI, MMF, Steroids | 236,772 (64.6%) |
| CNI, MMF | 62,667 (17.1%) |
| CNI, Steroids | 16,108 (4.4%) |
| CNI only | 2,673 (0.7%) |
| No CNI | 28,225 (7.7%) |
| Missing data | 19,959 (5.5%) |
| **Delayed Graft Function** |  |
| No | 297,564 (81.2%) |
| Yes | 65,778 (17.9%) |
| Missing data | 3,062 (0.8%) |
| **Previous Organ Transplant** |  |
| No | 319,526 (87.2%) |
| Yes | 23,134 (12.8%) |
| **Dialysis Before Transplant** |  |
| No | 59,063 (16.1%) |
| Yes | 296,348 (8.9%) |
| Missing data | 10,993 (3.0%) |

**BMI:** Body Mass Index; **CNI:** Calcineurin Inhibitor; **CIT:** Cold Ischemia Time; **DD:** Deceased Donor; **ECD:** Expanded Criteria Donor; **MMF:** Mycophenolate Mofetil; **PRA:** Panel Reactive Antibodies. Data are shown as the mean with standard errors for continuous variables. Categorical variables are reported as percentages.

Figure 2, **Panels A and B** show the mean IGF-1 levels ± 2SD values for males and females by age in the reference cohort. The peak values of IGF-1 were at age 15 in both sexes. Figure 2**, Panel C** shows that the mean of pooled IGF-1 levels for males and females increases approximately six-fold from birth, peaking at 15-years and subsequently diminishing into older age. **Panel D** shows the incident DCGF rate by the age at which the recipient underwent KTx illustrating parallel age-related changes in IGF-1 and incident DCGF rates. **Panel E** shows the incident DCGF rate at 1-year (short-term), 3-year (intermediate-term), and 10-year (long-term) outcomes illustrating that while at 1-year there is no detectable increase in DCGF in the 15-yr age group, at the intermediate and especially at long-term (10-year) follow-up the relationship between IGF-1 level by age and DCGF rates by age appear remarkably similar. **Panel F** uses the same data but shows the time to median allograft survival to illustrate the magnitude of the changes observed wherein a 15-year-old recipient has a median graft survival of only 12 years in contrast to a 60-year-old recipient whose median graft survival approaches 20-years. The joint analysis of the OPTN and Bidlingmaier studies thus shows that intermediate and long-term DCGF risk, but not short-term risk, is highest when circulating IGF-1 levels are highest and *vice versa*.

**Figure 2.**
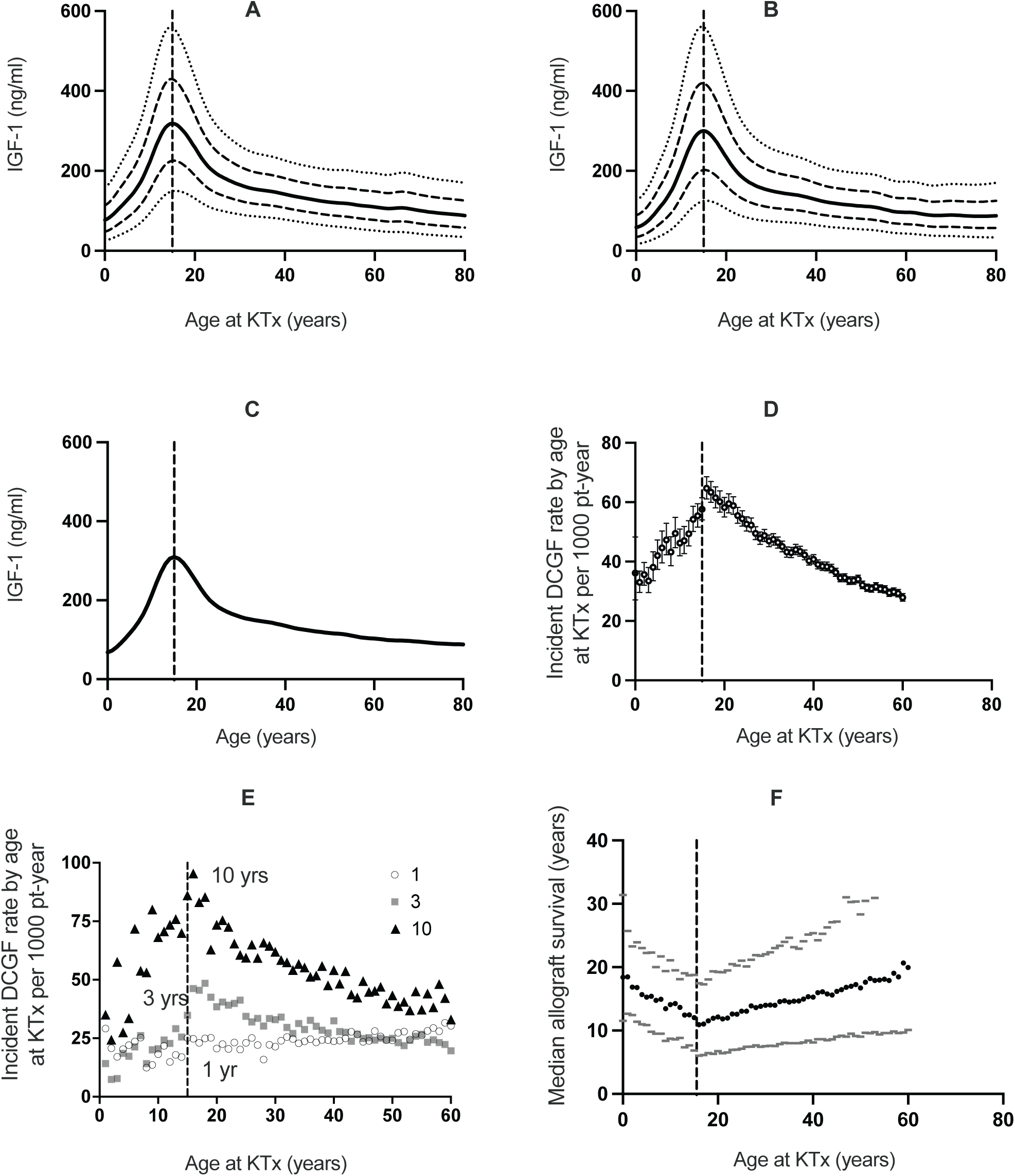
Legend: Comparison between two reference populations where IGF-1 levels and transplant outcomes by age are tested. **Panel A:** Age-specific average (solid dark line) IGF-1 levels from Bidlingmaier et al. for males, with their 1 SD (dashed lines) above and below mean values. Dotted lines represent the 2 SD values. **Panel B:** Age-specific average (solid grey line) IGF-1 levels for females, with 1 SD (dashed lines) above and below mean values. Dotted lines represent the 2 SD values. **Panel C:** Solid line shows mean IGF-1 levels for pooled males and females, which are not different, both peaking at 15 years of age. **Panel D:** Incident DCGF rate per 1000 patient-years with 95% CI. **Panel E:** Breakdown of incident DCGF rate per 1000 patient years into short-term (1 year), intermediate-term (3 years), and long-term (10 years) outcomes. **Panel F:** Median death censored allograft survival rates in dark circles along with their 25% and 75% values in grey. The dashed vertical line represents 15 years of age across all panels.

### Cohort 2: Clinical Study

To obtain more granular data including measured pretransplant IGF-1 levels, we conducted a clinical study of 216 patients whose sample size was determined by a pilot study (see methods). The clinical study tested the relationship of early allograft IGF-1 exposure on DCGF as the primary outcome, high-grade proteinuria (>1g/g creatinine) as a secondary outcome, and rejection as an exploratory outcome. The cohort had a median follow-up of 8.6 years. **Table 2** details the characteristics of donors and recipients of this cohort. The mean age of recipients was 47.4 ± 17.7 years, spanning from 1.8 to 77 years. A single circulating IGF-1 level has shown utility over pulsatile GH levels given the very low coefficient of variation noted in IGF-1 levels noted in multiple studies(36–39). Serum IGF-1 levels ranged from 16.3 to 624.6 ng/ml, averaging 206.8 ± 114.2 ng/ml. The average serum IGF-2 levels were 1067.6 ± 565.8, ranging from 144-2655.6 ng/ml. Figure 3A demonstrates the relationship between IGF-1 levels by age at KTx and while Figure 3B shows the relationship between IGF-2 levels by age at KTx both of which are very different from those seen with IGF-1 despite the significant correlation observed between IGF-1 and IGF-2 levels as shown in Figure 3C (r=0.48, p<0.0001)

**Figure 3:**
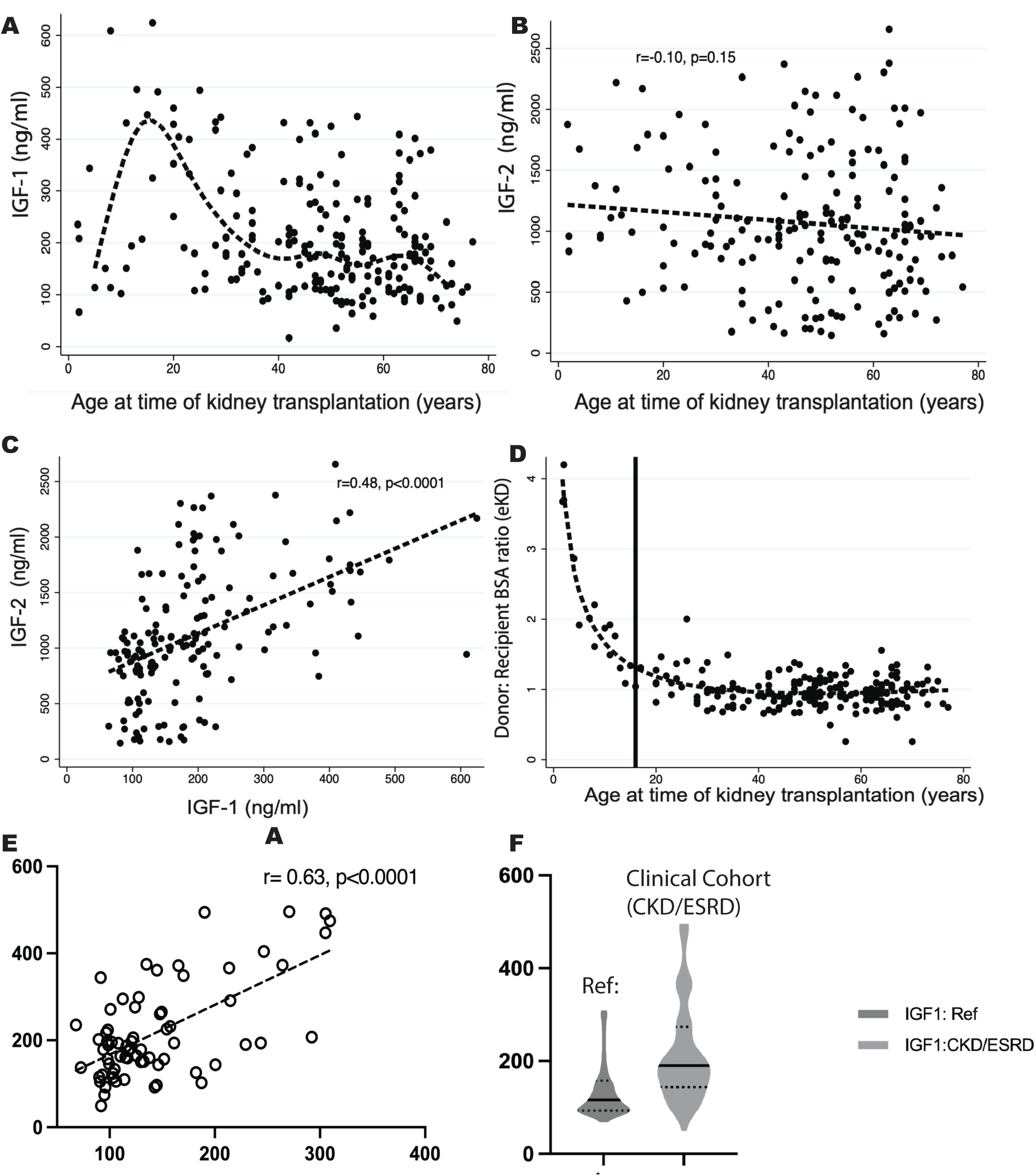
**Panel A:** Relationship between pre-transplant IGF-1 levels and recipient age at kidney transplantation. A wide range of IGF-1 levels was observed across all ages, but the averaged curve is similar to that of the reference population (shown in Figure 2C). **Panel B:**Relationship between pre-transplant IGF-2 levels and recipient age. A wide range of IGF-2 levels was observed across all ages. **Panel C:**A correlation between pre-transplant IGF-1 and IGF-2 levels was observed (r = 0.48, p < 0.0001). Note that IGF-2 was below the level of detection in 27 samples (12.5%). **Panel D**: Illustrates the relationship between the donor:recipient BSA ratio and recipient age at kidney transplantation. This relationship is similar to that found in the OPTN dataset (not shown). The vertical line denotes age 15 years. **Panel E:** Correlation plot between measured average IGF-1 levels by age in the clinical cohort of CKD and ESKD patients before KTx versus average IGF-1 levels by age in the Bidlingmaier et al. study. **Panel F:** Distribution of average IGF-1 values by age in the healthy reference population compared to the clinical cohort of CKD and ESKD patients. Although a direct comparison should not be made, as supported by the literature, IGF-1 levels are higher in CKD and ESKD patients.

**Table 2:**
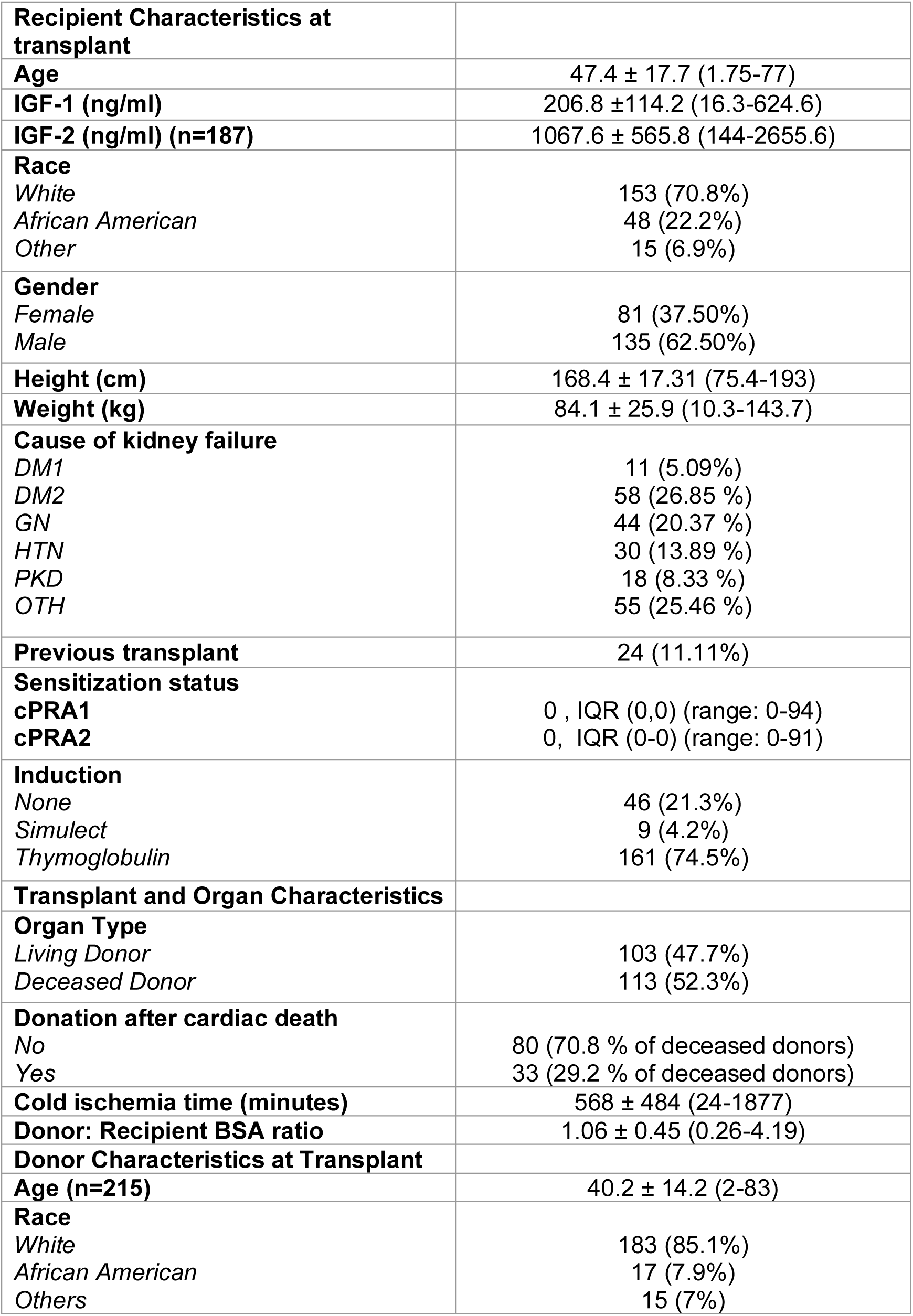
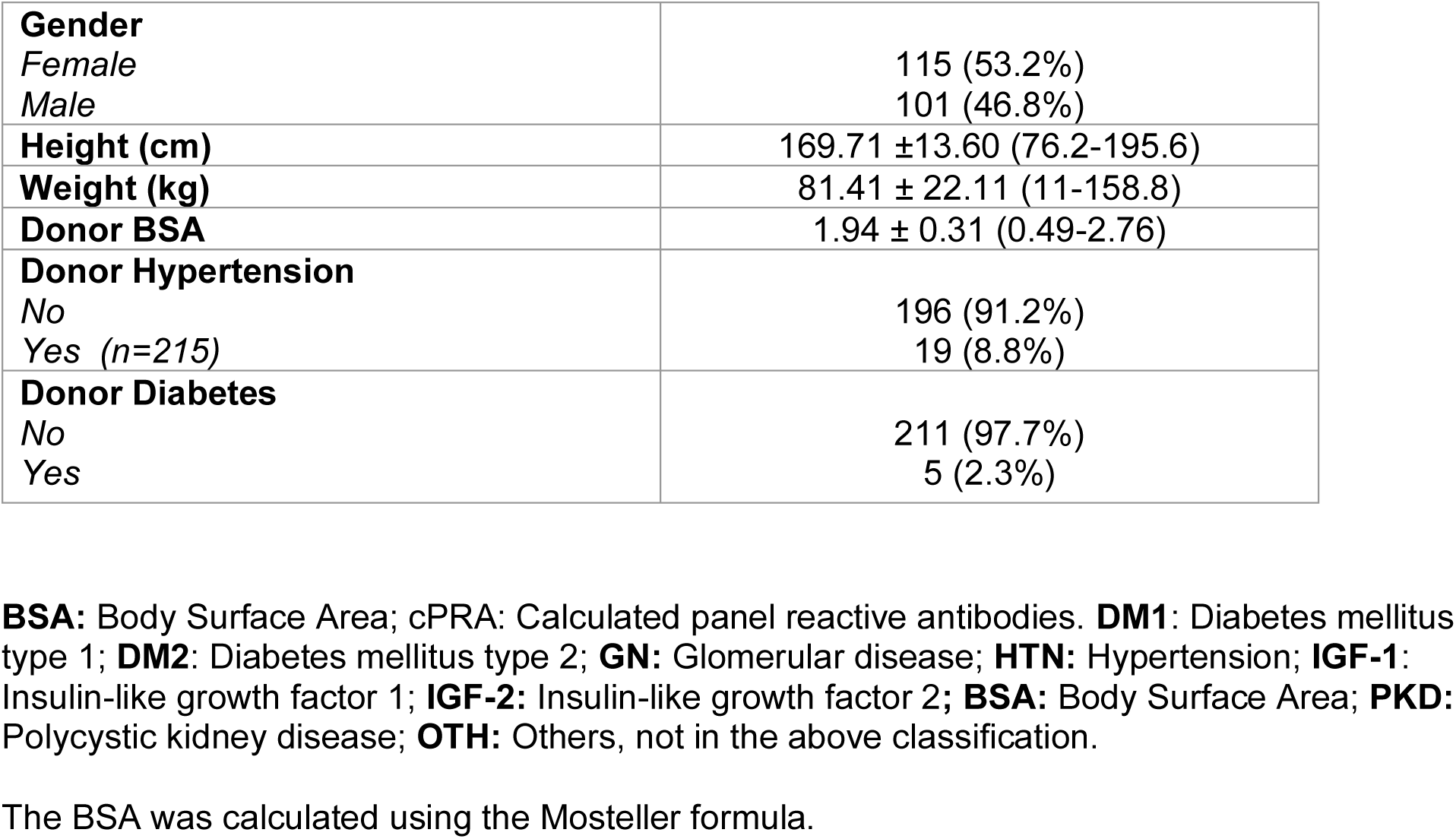
Donor, recipient, and transplant characteristics of the University of Michigan cohort used for analysis (n=216).

|  |  |
| --- | --- |
| **Recipient Characteristics at transplant** |  |
| **Age** | 47.4 ± 17.7 (1.75-77) |
| **IGF-1 (ng/ml)** | 206.8 ± 114.2 (16.3-624.6) |
| **IGF-2 (ng/ml) (n=187)** | 1067.6 ± 565.8 (144-2655.6) |
| **Race** |  |
| *White* | 153 (70.8%) |
| *African American* | 48 (22.2%) |
| *Other* | 15 (6.9%) |
| **Gender** |  |
| *Female* | 81 (37.50%) |
| *Male* | 135 (62.50%) |
| **Height (cm)** | 168.4 ± 17.31 (75.4-193) |
| **Weight (kg)** | 84.1 ± 25.9 (10.3-143.7) |
| **Cause of kidney failure** |  |
| *DM1* | 11 (5.09%) |
| *DM2* | 58 (26.85 %) |
| *GN* | 44 (20.37 %) |
| *HTN* | 30 (13.89 %) |
| *PKD* | 18 (8.33 %) |
| *OTH* | 55 (25.46 %) |
| **Previous transplant** | 24 (11.11%) |
| **Sensitization status** |  |
| **cPRA1** | 0 , IQR (0,0) (range: 0-94) |
| **cPRA2** | 0, IQR (0-0) (range: 0-91) |
| **Induction** |  |
| *None* | 46 (21.3%) |
| *Simulect* | 9 (4.2%) |
| *Thymoglobulin* | 161 (74.5%) |
| **Transplant and Organ Characteristics** |  |
| **Organ Type** |  |
| *Living Donor* | 103 (47.7%) |
| *Deceased Donor* | 113 (52.3%) |
| **Donation after cardiac death** |  |
| *No* | 80 (70.8 % of deceased donors) |
| *Yes* | 33 (29.2 % of deceased donors) |
| **Cold ischemia time (minutes)** | 568 ± 484 (24-1877) |
| **Donor: Recipient BSA ratio** | 1.06 ± 0.45 (0.26-4.19) |
| **Donor Characteristics at Transplant** |  |
| **Age (n=215)** | 40.2 ± 14.2 (2-83) |
| **Race** |  |
| *White* | 183 (85.1%) |
| *African American* | 17 (7.9%) |
| *Others* | 15 (7%) |
| **Gender** |  |
| Female | 115 (53.2%) |
| Male | 101 (46.8%) |
| **Height (cm)** | 169.71 ±13.60 (76.2-195.6) |
| **Weight (kg)** | 81.41 ± 22.11 (11-158.8) |
| **Donor BSA** | 1.94 ± 0.31 (0.49-2.76) |
| **Donor Hypertension** |  |
| No | 196 (91.2%) |
| Yes (n=215) | 19 (8.8%) |
| **Donor Diabetes** |  |
| No | 211 (97.7%) |
| Yes | 5 (2.3%) |

**BSA:** Body Surface Area; cPRA: Calculated panel reactive antibodies. **DM1:** Diabetes mellitus type 1; **DM2:** Diabetes mellitus type 2; **GN:** Glomerular disease; **HTN:** Hypertension; **IGF-1:** Insulin-like growth factor 1; **IGF-2:** Insulin-like growth factor 2; **BSA:** Body Surface Area; **PKD:** Polycystic kidney disease; **OTH:** Others, not in the above classification.
The BSA was calculated using the Mosteller formula.

Figure 4 provides a simplified visual of the overall hypothesis for how amount of allograft IGF-1 exposure was defined by leveraging the recipient’s circulating IGF-1 levels at time of transplantation and the degree of hyperfiltration experienced by that graft. The effective kidney dose (eKD), determined by the donor:recipient BSA ratio, determines amount of hyperfiltration(8). **Table 3** summarizes the association between allograft IGF-1 exposure (IGF-1*eKD) with outcomes, including DCGF, proteinuria, and BPAR, and additional analysis for TCMR. As modeled in Figure 5 the interaction effect is most obvious at values lower than the mean eKD in our cohort, such that for an individual recipient with higher IGF-1 levels, an eKD of less than 50% (less than one-kidney equivalent) is associated with increased risk of DCGF, proteinuria, and TCMR risk. Conversely, an eKD >50% (more than one kidney equivalent) tended to protect against all outcomes examined.

**Figure 4:**
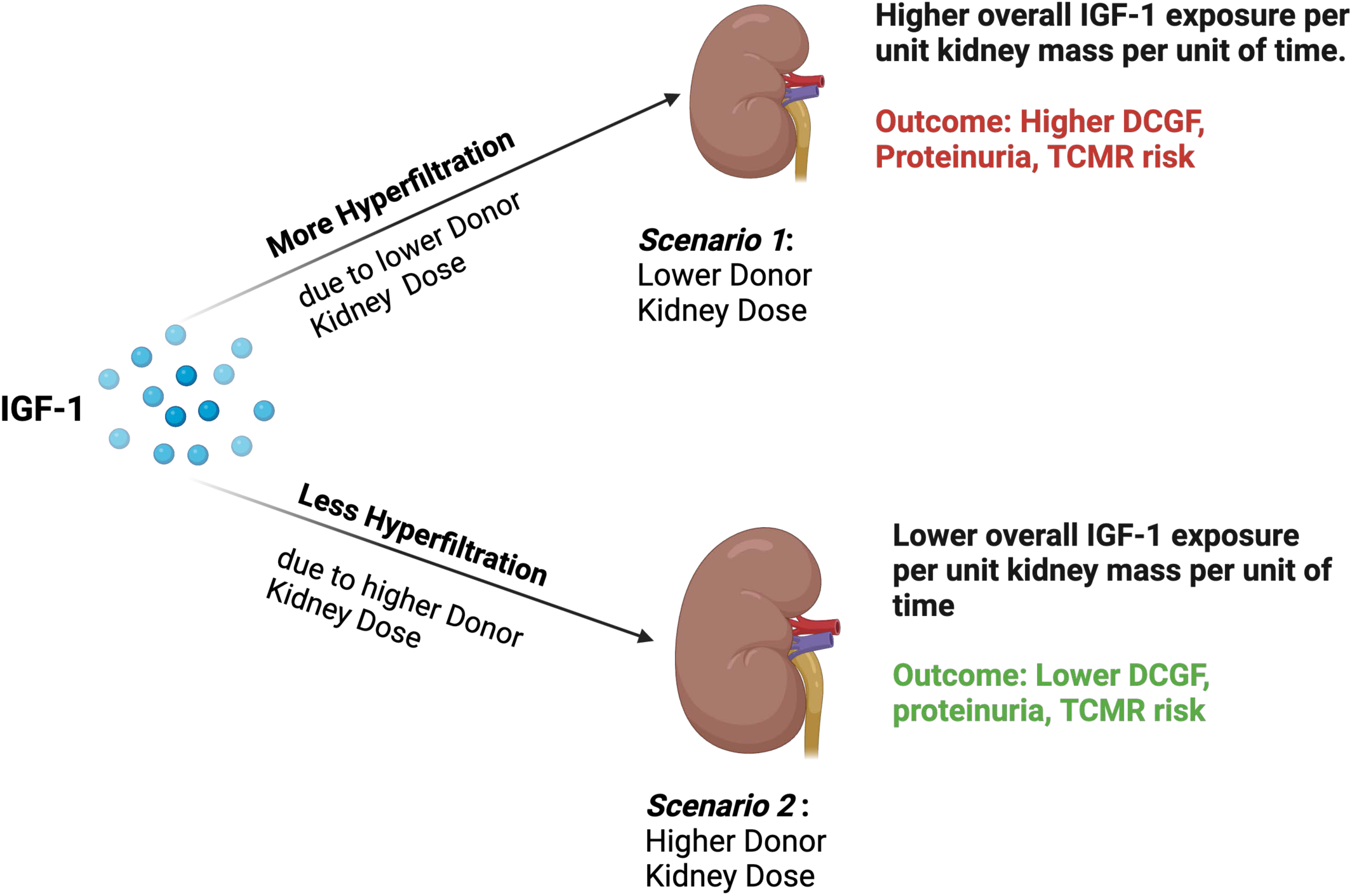
Diagrammatic illustration of how IGF-1 exposure was defined leveraging the circulating IGF-1 levels in the recipient, with the kidney dose determining hyperfilteration and thus overall IGF-1 exposure. At a given IGF-1 level, a lower kidney dose is expected to be accompanied by increased hyperfiltration, which leads to a higher IGF-1 delivery to the allograft. The increase in IGF-1 delivery alone or in combination with the lower functional mass is expected to increase overall IGF-1 exposure per unit of donor kidney dose (per unit of time, given blood flow rate is a function of time) driving inferior allograft survival. Conversely, at higher kidney doses, the extent of hyperfiltration is lower, which leads to reduced overall IGF-1 exposure per unit of kidney mass (per unit of time) leading to superior allograft survival.

**Figure 5:**
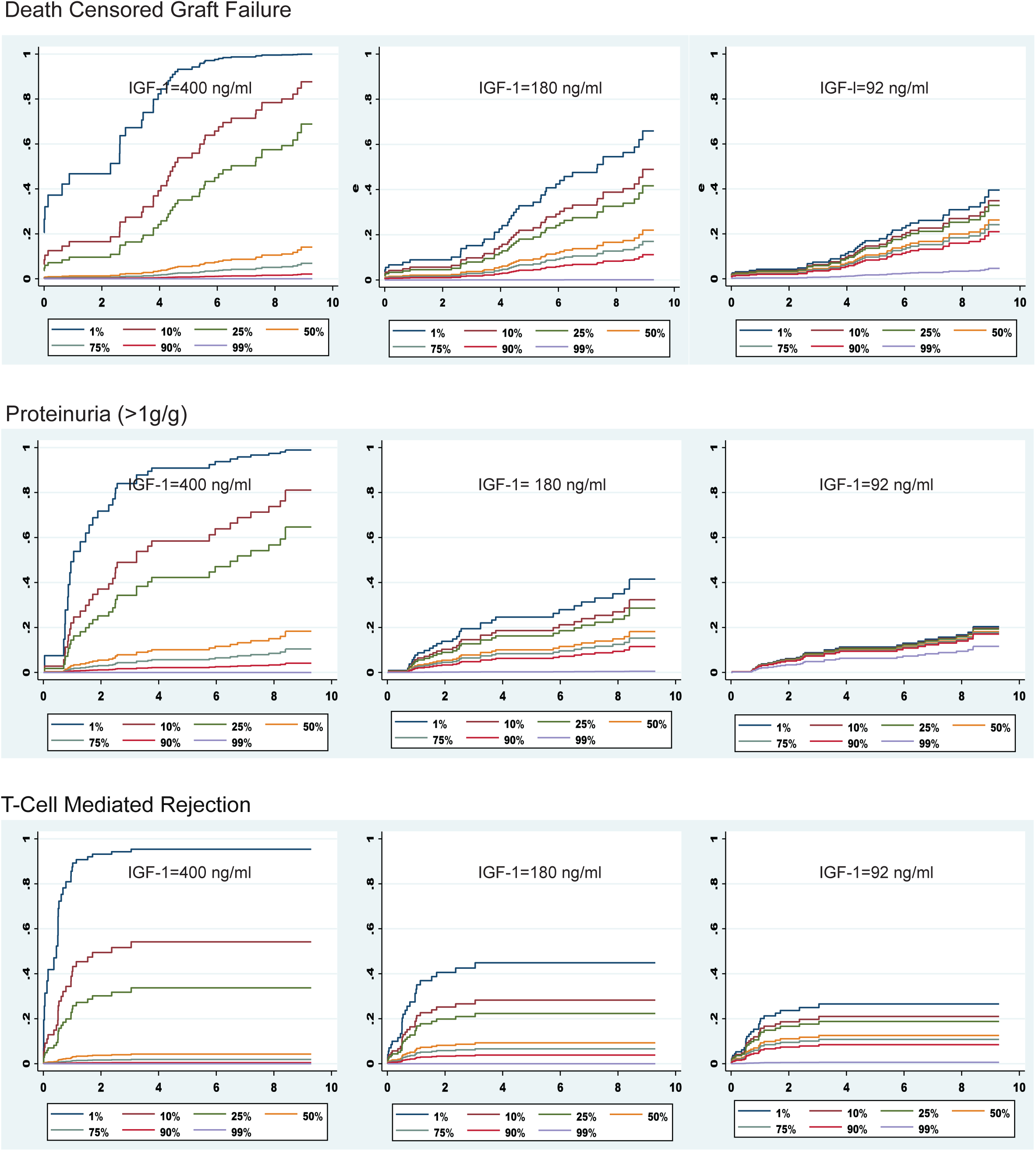
Relationship between IGF-1 levels and outcomes of interest in relation to the estimated kidney dose (eKD) delivered to the recipient at Kidney Transplantation (n=216). The survival curves help visualize covariate-adjusted outcomes from the multivariable Cox regression model. Survival curves are generated across different IGF-1 values (90th, 50th, and 10th percentiles of the distribution in the clinical cohort) and eKD values (1st, 10th, 25th, 50th, 75th, 90th, and 99th percentiles corresponding to 0.25, 0.37, 0.42, ^∼^0.50, 0.56, 0.67, 1.83 kidney equivalents). Mean centering was performed for the eKD variable to allow for interpretation of the direct effect of IGF-1 at the mean of eKD in our clinical cohort. For DCGF and proteinuria, the Cox regression models were adjusted for donor age, donor race, donor gender, donor type (living or deceased), donor body surface area, recipient age, recipient race, recipient gender, and cold ischemia time. For the TCMR (and separately for BPAR), in addition to the above variables, adjustments were also made for pretransplant calculated panel reactive antibodies class 1 or 2 and the number of previous transplants

**Table 3:** Relationship between IGF-1 exposure (IGF-1*eKD interaction) and Death Censored Graft Failure (Primary Outcome), Proteinuria and BPAR (Secondary Outcome), and TCMR as an additional sensitivity analysis. The table shows **selected variables** from a multivariable Cox regression model for death-censored graft failure, proteinuria, Biopsy Proven Acute Rejection (BPAR), and T-cell mediated Rejection. The DCGF models include donor age, donor race, donor gender, donor body surface area, type of donor (living versus deceased), cold ischemia time in minutes, recipient age, recipient race, and recipient gender. The secondary outcomes of BPAR and the sensitivity analysis of TCMR, in addition to the above variables, were also adjusted for pretransplant calculated panel reactive antibody class 1 and 2 and the number of previous transplants to adjust for baseline sensitization status at the time of kidney transplantation. Supplemental Tables 1-7 show the entire multivariable Cox models.

| Variables | Haz. ratio | Std. err. | P value | LCL | UCL |
| --- | --- | --- | --- | --- | --- |
| **DCGF** |  |  |  |  |  |
| **IGF-1\*eKD** | 0.985 | 0.005 | 0.007 | 0.974 | 0.996 |
| IGF-1 | 0.844 | 0.087 | 0.099 | 0.690 | 1.033 |
| eKD | 1.014 | 0.015 | 0.334 | 0.986 | 1.043 |
| **Proteinuria (>1 g/g)** |  |  |  |  |  |
| **IGF-1\*eKD** | 0.987 | 0.005 | 0.013 | 0.976 | 0.997 |
| IGF-1 | 0.952 | 0.091 | 0.607 | 0.789 | 1.149 |
| eKD | 1.021 | 0.016 | 0.186 | 0.990 | 1.053 |
| **BPAR** |  |  |  |  |  |
| **IGF1\*eKD** | 0.994 | 0.003 | 0.068 | 0.987 | 1.000 |
| IGF-1 | 0.881 | 0.074 | 0.133 | 0.748 | 1.039 |
| eKD | 1.006 | 0.012 | 0.599 | 0.983 | 1.030 |
| **TCMR** |  |  |  |  |  |
| **IGF-1\*eKD** | 0.985 | 0.007 | 0.044 | 0.970 | 1.000 |
| IGF-1 | 0.787 | 0.098 | 0.055 | 0.616 | 1.005 |
| eKD | 1.005 | 0.020 | 0.791 | 0.968 | 1.044 |

#### Death-Censored Graft Failure (DCGF)

Forty-four of the 216 patients experienced DCGF by a median of 8.6 years of follow-up. IGF-1 exposure (IGF-1*eKD) was significantly associated with DCGF (p<0.01) (**Table 2**). Figure 5 models the association of high IGF-1 levels and a smaller kidney dose (thus higher IGF-1 exposure for the kidney dose delivered) on DCGF risk (Figure 5**, top panels**). Higher IGF-1 exposure was associated with increased DCGF risk. Conversely, a larger kidney dose (lower IGF-1 exposure) decreased DCGF risk. Notably, at lower IGF-1 levels, kidney dose is predicted to be less consequential on DCGF risk. For the complete multivariable Cox models, refer to Supplementary Table S1. Additional sensitivity analyses, including adjustment of pretransplant sensitization (Supplementary Table S2), development of *de novo* donor-specific antibodies (*dnDSA*) after KTx (Supplementary Table S3), and BPAR (Supplementary Table S4). IGF-1 levels alone or eKD alone were not statistically significantly associated with DCGF at the mean eKD values in our cohort, suggesting that the effect of IGF-1 levels on DCGF is modulated by the amount of kidney dose delivered at KTx.

#### Proteinuria (>1g/g creatinine in urine)

Proteinuria is a well-established surrogate for long-term allograft outcomes(40).Thirty-three of the 216 patients developed proteinuria >1g/g by a median of 2.4 years post-KTx. IGF-1 exposure (IGF-1*eKD) was significantly associated with proteinuria (p<0.01) (**Table 2**). Figure 5**, middle panels**, models the effect of IGF-1 exposure on proteinuria. Like DCGF, higher IGF-1 exposure is associated with increased proteinuria, while this effect is less pronounced at lower IGF-1 exposure. For the complete multivariable Cox models, see Supplementary Table S5.

#### Biopsy Proven Acute Rejection (BPAR)

Forty-four patients were identified as experiencing BPAR from routine surveillance biopsies at 3, 6, and 12 months and clinically indicated biopsies.

The median time to any rejection was 0.95 years, with the median times for TCMR at 0.54 years, antibody-mediated rejection at 2.9 years, and mixed rejection at 3.1 years. In the sensitivity analysis, higher IGF-1 exposure (IGF-1*eKD) was associated with an increased tendency towards BPAR (p=0.07) (**Table 2**) and a significant association with TCMR (p=0.04). Figure 5, **bottom panels** model the effect of IGF-1 exposure on TCMR risk. For the complete multivariable Cox models, see Supplementary Tables S7. There was no significant relationship detected between pre-transplant IGF-2 levels on DCGF, proteinuria, or alloimmune responses (data not shown).

### Cohort 3: Genotype Study

The SNP variant (*rs35767*) in the regulatory region of the *IGF1* gene is associated with elevated circulating IGF-1 levels(33–35,41). A previous study has noted that genetic factors account for 38% of the variance in IGF-1 levels(42). Thus, if our hypothesis is correct, a genetic variant in IGF-1 that increases the levels of circulating IGF-1 in the recipient would also be expected to be associated with inferior long-term outcomes. A multivariable analysis of the National Institute of Health-funded Genomics of Chronic Allograft Rejection (GoCAR) (n=527) and Clinical Trials in Organ Transplantation (CTOT) (n=197) cohorts, supplemented by a meta-analysis pooling these studies (n=724) depicted in Figure 6 (43–46). These data suggest that possessing even one risk allele is associated with a 50% (HR =1.50, p = 0.01) increase in the risk of DCGF. These data further support the hypothesized relationship between IGF-1 exposure and DCGF. **Table 4** shows the multivariable analysis to explore the effect of presence of risk allele *rs35767* on recipient DCGF risk.

**Figure 6:**
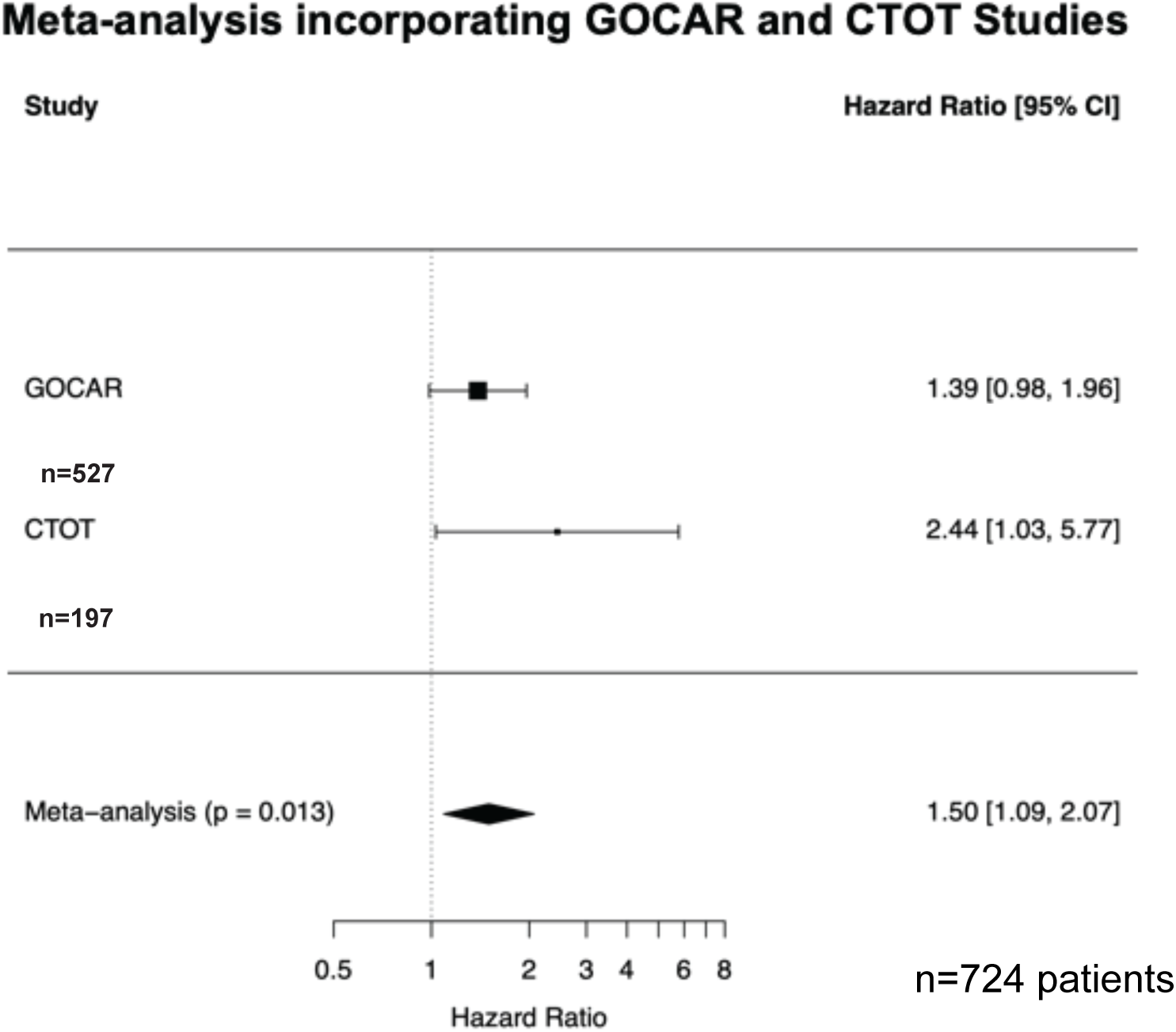
A meta-analysis investigating the relationship between a Single Nucleotide Polymorphism (SNP) in the IGF-1 gene, rs35767, and Kidney Allograft Failure. The meta-analysis uses data from the Genomics of Chronic Allograft Rejection (GoCAR) and the Clinical Trials in Organ Transplantation (CTOT) 01/17 studies. In the GoCAR study, the risk of DCGF, with an adjusted hazards ratio (aHR) of 1.38, did not reach clinical significance (p = 0.06). In contrast, in the CTOT study, the aHR was 2.44, which achieved statistical significance (p = 0.001). The meta-analysis included 527 kidney transplant recipients from the GoCAR study and 197 patients from the CTOT study and was significant (p = 0.013). There was a 50% increase in the risk of DCGF associated with recipient carriage of a single rs35767 risk allele (aHR = 1.50, p = 0.01).

**Table 4:** Risk of recipient death censored graft failure in the presence of one risk allele of SNP *rs35767*.

|  | 95% confidence interval | Hazard ratio | P value |
| --- | --- | --- | --- |
| **GoCAR (N = 527, with 197 having the risk allele), 73 DCGL events** |  |  |  |
| **rs35767<sup>a</sup>** | (0.98, 1.96) | 1.39 | 0.062 |
| **Recipient age** | (0.97, 1.01) | 0.989 | 0.249 |
| **Recipient weight** | (0.99, 1.01) | 0.998 | 0.731 |
| **Donor age** | (0.99, 1.03) | 1.009 | 0.318 |
| **HLA mismatch** | (1.03, 1.77) | 1.348 | 0.031 |
| **CTOT01/17 (N = 192, with 84 having the risk allele), 14 DCGL events** |  |  |  |
| **rs35767<sup>a</sup>** | (1.03, 5.77) | 2.44 | 0.042 |
| **Donor age** | (0.98, 1.07) | 1.023 | 0.338 |
| **Recipient weight** | (0.98, 1.00) | 0.99 | 0.168 |
| **HLA mismatch** | (1.03, 2.21) | 1.506 | 0.036 |
| **Recipient age** | (0.98, 1.05) | 1.013 | 0.499 |

<sup>a</sup>. Additive model with risk allele numbers 0, 1, 2

### Cohort 4: Analysis of IGF-1 pathway transcripts in Healthy and Transplanted Kidneys

Next, we analyzed cell-type specific transcriptional activity of kidney cells using single-cell RNA sequencing from healthy donor biopsies pre-implantation and surveillance biopsies from allograft recipients in the first post-transplant year allografts in which protocol kidney biopsies showed no pathologic abnormality(30,47). To investigate cell-specific IGF1 expression before- and early post-transplant, we focused on transcripts in the GH/IGF-1/IGF-1R pathway in these analyses. As depicted in Figure 7, *IGF1* transcript was predominantly expressed in podocytes and fibroblasts, with other kidney cell types showing minimal detectable expression. Podocytes also exhibited high *GHR* transcript levels. *GH* transcript was not detected in any kidney cell type (data not shown). *IGF1R* and *INSR* transcript were broadly expressed, particularly in podocytes and specific proximal tubular cells, reflecting their roles as gene reduplicates and involvement in forming hybrid receptors responsive to IGF-1(48). Notably, *IGFBP 1-6* expression tended to be cell-specific, possibly underscoring a refined regulatory mechanism within specific renal cell types(49). Interestingly comparisons between healthy preimplant kidneys and stable first-year kidney allografts with no histological abnormalities revealed no significant differences in *IGF1* transcript levels (Figure 8A), but significantly decreased *IGF1R* and *GHR* transcripts (Figure 8B), akin to findings from rodent uni-nephrectomy models(28,50). These data are consistent with the hypothesis that the normal "compensatory" adaptive response of the allograft is to down-regulate its intrinsic IGF-1 pathway machinery in response to high level extrinsic IGF-1 exposure from the circulation as a result of hyperfiltration.

**Figure 7:**
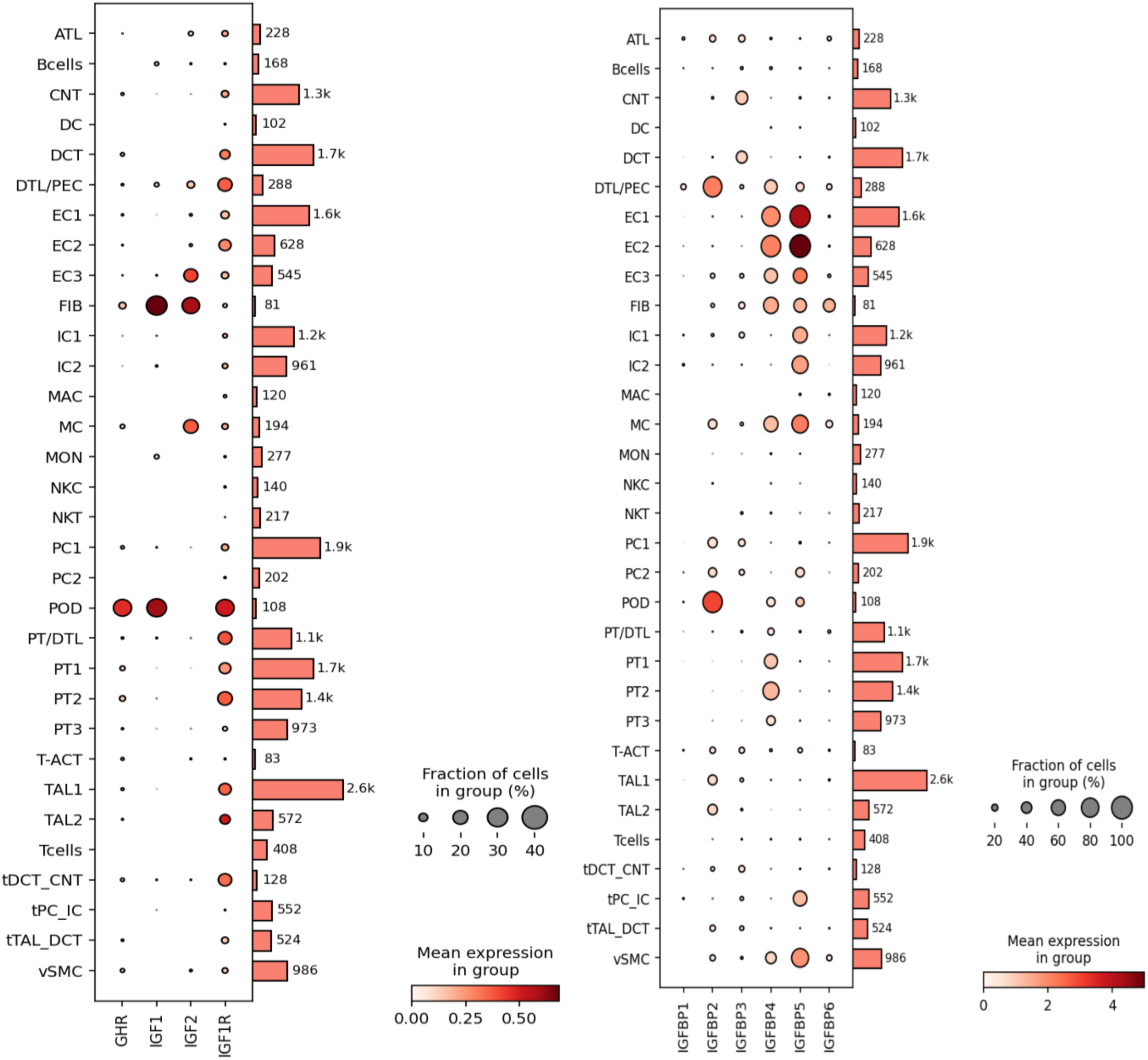
Single-cell transcriptomic analysis of IGF-pathway transcripts in kidney cells derived from biopsies of normal kidneys obtained prior to transplantation. Data are sourced from a previously reported dataset, wherein the criteria for cell identification are detailed (Ref. 41). The number of cells available for analysis for each cell type is displayed to the right of each panel. The cell types are listed alphabetically from top to bottom (definitions below). For each transcript, the dot size represents the proportion of cells within the cell type that expressed detectable transcripts, while the intensity of the dot color reflects the average amount of transcript detected across all cells. The dot size and intensity calibration are shown at the lower right of each panel. The panels display transcripts for Insulin-like Growth Factor-1 (IGF1), IGF-1 Receptor (IGF1R), Growth Hormone Receptor (GHR), Insulin-like Growth Factor-2 (IGF2), and Insulin Receptor (INSR); as well as Insulin-like Growth Factor Binding Proteins 1-6 (IGFBP1-6). Cell designations are as follows: ATL (Ascending Thin Limb), BCells (B Cells), CNT (Connecting Tubule), DC (Dendritic Cells), DCT (Distal Convoluted Tubule), DTL (Descending Thin Limb), EC1 (Arteriolar Endothelial Cells), EC2 (Glomerular Endothelial Cells), EC3 (Peritubular Capillaries), FIB (Fibroblasts), IC1/2 (Intercalated Cells 1 and 2), MAC (Macrophages), MC (Mesangial Cells), MON (Monocytes), NKC (Natural Killer Cells), NKT (Natural Killer T Cells), PC1/2 (Principal Cells Cluster 1 and 2), POD (Podocytes), PT/DTL (Proximal Tubule/Descending Thin Limb), PT1/2/3 (Proximal Tubule Sub-clusters 1, 2, and 3), T-ACT (Activated T Cells), TAL1/2 (Thick Ascending Limb 1 or 2 Sub-cluster), TCells (T Cells), tDCT_CNT (Transitional cells of DCT with CNT), tPC_IC (Transitional Principal and Intercalated Cells), tDAL_DCT (Transitional cells between TAL and DCT), vSMC (Vascular Smooth Muscle Cells).

**Figure 8:**
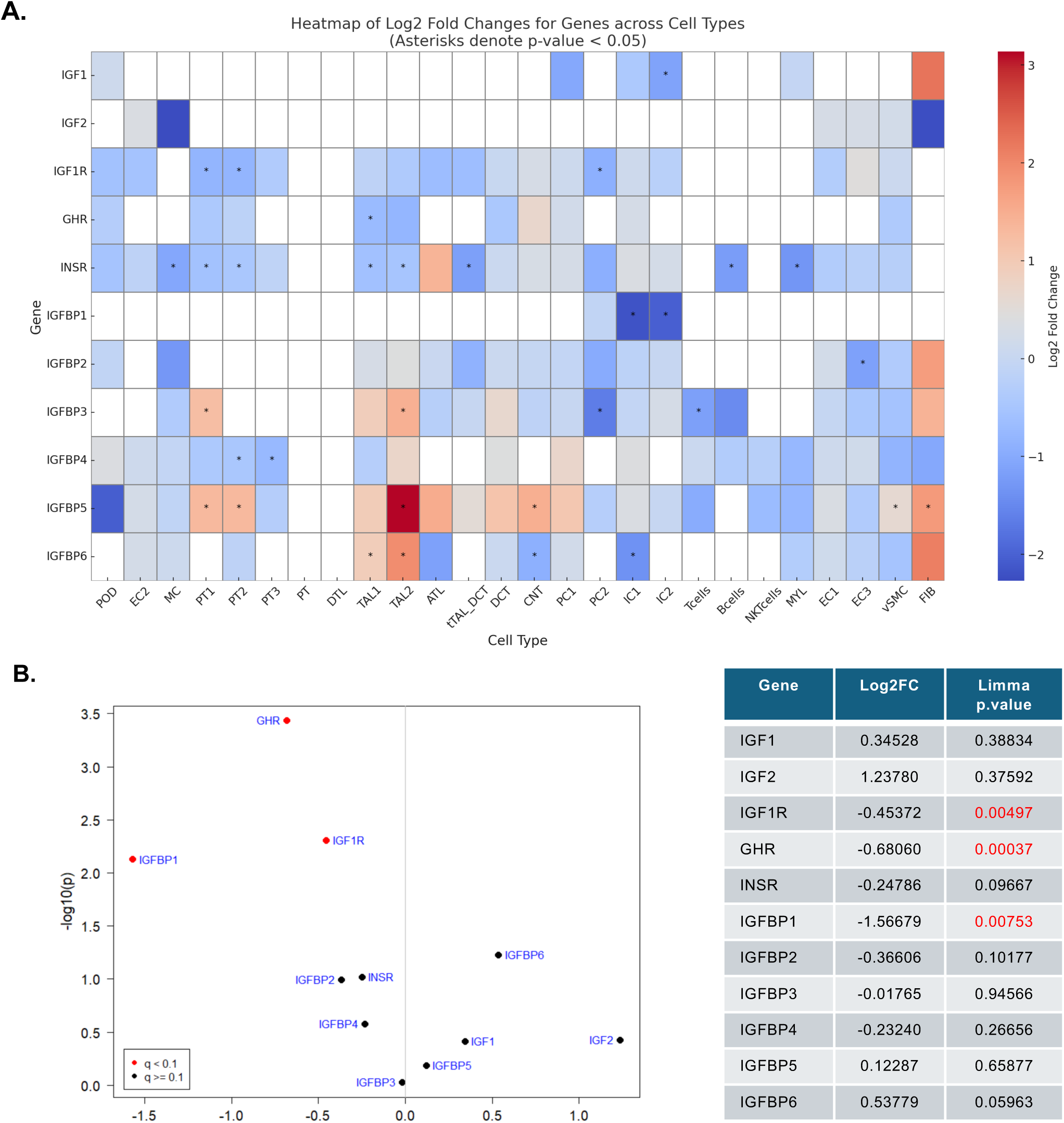
**Panel A:** Heatmap of key genes in the GH-IGF1 pathway at a cell-specific level. The heatmap was created using pseudo-bulking of cell-level gene expression profiles between normal kidney tissue from living donors in a bi-nephric state versus solitary kidney allografts. (Ref. 41) A negative log2 fold change, represented by blue, indicates a relative reduction in the expression of specific genes in first-year kidney allografts (n=14) compared to a bi-nephric, healthy human control group (n=20). Conversely, a positive log2 fold change, indicated by red, signifies increased gene transcript expression. The absence of color denotes that the transcript was not detected in the analysis. An asterisk (*) marks genes for which the log2 fold change differences between the two groups reached statistical significance (P < 0.05). **Panel B:** Volcano plot depicting the differential expression analysis of selected genes within the IGF-1 signaling pathway, using transplant kidney pseudo-bulk RNA sequencing. This analysis contrasts gene expression in first-year kidney allografts with that in bi-nephric healthy individuals. In the plot, individual genes are represented by points, plotted according to their fold change (x-axis) and the negative logarithm (base 10) of their p-value (y-axis). The color of each point indicates the significance level of the associated p-value, with red denoting very high significance

## Discussion

Our multicohort studies support the hypothesis that KTx longevity is associated with IGF-1 exposure from the recipients circulation that occurs early after KTx. A population-level study identified that patients undergoing KTx in adolescence and early adulthood, when ambient GH-IGF-1 levels are highest, have the highest intermediate and long-term DCGF rates, while lower IGF-1 levels at extremes of age were associated with lower DCGF rates. The lower DCGF rates translate to a difference in median allograft longevity of about 12 years for a 15-year old versus 19.9 years for a 60 year old recipient. A higher IGF-1 exposure was associated with a higher risk of DCGF (primary outcome) and proteinuria (secondary outcome). Conversely, lower IGF-1 exposure was associated with a lower risk for DCGF and proteinuria. Notably, while not powered for immune-mediated effects, our exploratory analysis revealed an increased risk of TCMR associated with higher IGF-1 exposure and, conversely, a lower TCMR risk with lower exposure. The association between allograft IGF-1 exposure and outcomes was further buttressed by genotype studies using three NIAID KTx cohorts wherein an *IGF1* SNP linked to high circulating IGF-1 levels was associated with a higher DCGF risk(33–35). Finally, in alignment with prior Uni-Nx studies, our transcriptomic data supports the hypothesis that there is no net increase in *IGF1* transcript in first-year allografts(28,50). *GHR* and *IGF1R* transcript were also downregulated, suggesting the presence of an inhibitory feedback loop to ward off excess extrinsic GH and IGF-1 exposure from the circulation. Taken together, we provide strong support for a previously unrecognized link between GH-IGF-1 axis-mediated signaling and kidney allograft failure.

The relationship of circulating IGF-1 level to DCGF risk depended on the kidney dose delivered at KTx. Kidney dose determines relative kidney blood flow and degree of hyperfiltration(8). At higher circulating IGF-1 levels and a smaller kidney dose, the allograft is predicted to experience higher IGF-1 exposure relative to the kidney dose delivered. This increase in IGF-1 exposure was associated with higher DCGF, proteinuria, and TCMR risk. Conversely, at low circulating IGF-1 levels, the donor kidney dose is predicted to be less consequential for graft outcomes, likely due to lower IGF-1 exposure regardless of the delivered kidney dose. This observation has potential implications for expanding the deceased donor kidney pool, as discussed below.

Advanced Chronic Kidney Disease (CKD) and End-Stage Kidney Disease (ESRD) are accompanied by high circulating GH levels although free IGF-1 levels are in the normal range(51–54). IGFBP3 protein, the major IGF-binding protein in blood, is increased in Chronic Kidney Disease (CKD), thereby accounting for the increase in total IGF-1 level (bound and free IGF-1) in CKD that was also noted in this current study (Figure 3F) (54). Acid Labile Subunit (ALS) levels remain in the normal range in CKD(54). Furthermore, IGF-1 production and its metabolic clearance rate are comparable in CKD and healthy individuals(54).

Podocytes have a high expression of *GHR, IGF1R,* and *IGF1* transcripts, suggesting the kidney also has intrinsic IGF-1 production and regulatory mechanisms, a finding consistent with the kidney being a net producer of IGF-1(55,56). It is, therefore, plausible that high GH exposure from hyperfiltration at the time of KTx would tend to activate GHR on the podocyte to drive podocyte production of IGF-1, and that the combined effects of GH and IGF-1 would promote mTORC1 activation to drive podocyte hypertrophy. We conjecture that the reduction in *GHR* by podocytes and *IGF1R* transcript expression observed across many cell types by 3-12 months after transplantation in allografts with no detectable pathologic abnormality reflects a broader adaptive response to protect against excess GH and IGF-1 exposure in hyperfiltering allografts. Similar findings of reduced *IGF1R* transcript expression in hyperfiltering adult rodent remnant kidney models have previously been reported(50). Thus, the net effects of hyperfiltered GH (that would tend to increase podocyte IGF-1 production) versus high allograft exposure to hyperfiltered IGF-1 from blood (that would tend to down-regulate intrinsic IGF-1R transcript expression) could balance out in the stable allograft resulting in the absence of a significant alteration in the IGF-1 transcript, as observed in our data.

Consistent with ongoing podocyte and glomerular injury, increased IGF-1 exposure was associated with proteinuria of >1g/g. IGF-1 signaling is critical for podocyte health and function(57). Accumulating data also suggests that GH can mediate podocyte injury(58–61). Thus, our data would be consistent with the hypothesis that hyperfiltration drives increased exposure of the allograft to circulating GH and IGF-1, which leads to increased engagement of their corresponding receptors (GHR and IGF-1R) on the podocyte, driving hypertrophy, injury, and stress leading to accelerated podocyte detachment in KTx that we have reported previously(11–13,30).

IGF-1 exposure was also associated with a trend towards increased TCMR Banff grade 1A or higher(62). This observation is consistent with a rodent model of allotransplantation, which showed that smaller kidney doses exhibited higher rejection rates and chronic allograft nephropathy(63,64). Furthermore, early reintroduction of additional kidney dose was reported to reverse rejection and mitigate chronic allograft nephropathy consistent with kidney dose being related to rejection and overall graft survival. The reduction in rejection when providing a larger kidney dose is consistent with the hypothesis that increasing the donor kidney dose would reduce hyperfiltration-induced allograft IGF-1 exposure, potentially diminishing allo-immune events and other processes that drive rejection. Given that IGF-1 is known to activate immune response(65–67), these preliminary observations underscore the need for more extensive, dedicated *in vitro* and in *in vivo* studies focusing on the immune aspects of IGF-1 signaling as also emphasized by studies of the immune effects of IGF-1R blockade in thyroid eye disease (68).

The genotype investigation revealed a robust association between the *rs35767* SNP and clinical outcomes, indicating that possessing even a single risk allele is associated with a 50% elevation in the risk of DCGF. This finding is not surprising because genetic variations account for 38% of the variance in IGF-1 levels(42). This variant’s minor allele frequency (MAF), as reported in the ALFA project (which consolidates allele frequencies across dbGaP), was A=0.18 (69). The MAF varied by population, with the highest frequency in African populations and Americans of immediate African ancestry (MAF=0.48) and the lowest frequency observed in individuals of European ancestry (MAF=0.16). Similar MAF was observed across self-reported ancestry in a CKD cohort (NEPTUNE). Supplementary Table S8 breaks down enrichment in this SNP by self-reported ancestry in the NEPTUNE cohort. Corroborating these findings, a recent report from Japan has also linked this genetic variant with worsening kidney function over the long term(33).These observations underscore the need for additional research to determine how this SNP influences kidney and allograft health, especially considering the high MAF for SNP *rs35767* and the pronounced health disparities including both CKD prevalence and worse allograft survival among the African American community in the United States(70,71).

Interpreting the role of IGF-1 in the allograft requires understanding the GH-IGF axis biology in the kidney, which has been the subject of extensive investigation and excellent reviews(55,72,73). The liver produces IGF-1 under the control of pituitary-derived GH. The kidney is also a net producer of IGF-1, with single-cell analysis studies demonstrating podocytes and fibroblasts as the major cell types transcribing IGF-1 (Figure 6)(56). In the circulation, free IGF-1 (7.5kDa) makes up only about 1% of total IGF-1 in blood. The remaining 99% of IGF-1 (and IGF-2) circulates bound to IGF-binding proteins (IGFBP1-6), particularly IGFBP3 and IGFBP5. Notably, IGFBPs are a family of gene-reduplicated proteins that bind IGF-1 with 10-100-fold higher affinity than the IGF-1R, thereby regulating local IGF-1 availability(74). IGFBPs release IGF-1 locally in response to proteolytic and stereochemical modulations induced by binding to matrix and other molecules(74–76). IGF-IGFBP bi-molecular complexes have molecular weights ranging from about 25-45kDa. The major IGF-BPs in the blood (IGFBP3 and IGFBP5) also bind to the ALS in blood to form trimolecular 150kDa complexes with a longer half-life(77,78). Both free IGF-1 and the IGF-IGFBP bi-molecular complexes would be expected to cross the normal glomerular filtration barrier to gain access to the urinary space, especially under hyperfiltration conditions. Increased renal blood flow would also increase IGF access to the kidney interstitial space and the basolateral surface of tubular cells via peritubular capillaries(79,80).

GH was not measured in the clinical study because it is released in a pulsatile and diurnal manner that makes random measurements of limited use. Thus, a single IGF-1 measurement is routinely used as a stable readout for net GH effects given the very low coefficient of variation in IGF-1 level(37–39,81). However, IGF-1 is released in response to GH acting on the GHR in the liver and other cells that express the GHR, including podocytes. GH and IGF-1 in combination can therefore both act via mTORC1 to amplify cell hypertrophy and hypertrophic stress. This is illustrated by murine models where over-expression of IGF-1 causes glomerular enlargement but not glomerulosclerosis, while over-expression of GH to cause the same level of IGF-1 causes both glomerular enlargement and glomerulosclerosis(82). To emphasize that the effects associated with IGF-1 may also be caused in part by GH we use the term “GH-IGF-1 axis” in this report.

IGF-2 levels in blood are generally 3-5-fold higher than IGF-1 levels(72). IGF-2 has similar binding to IGFBPs and ALS, and binds to and activates the IGF-1R(20–22). Therefore, higher pretransplant IGF-2 levels might also have been expected to be related to allograft longevity. However, we found no statistically significant relationship between IGF2 alone or its interaction with kidney dose driving any tested outcomes. Notably, in our rodent UniNx model, IGF-1 was preferentially hyperfiltered to appear in the urine compared to IGF-2, possibly due to differential binding to its carrier IGFBPs(27).

Medication non-compliance in the teenage years has previously been suggested as a cause of increased graft loss in this time period(83,84). Our population level analysis showed that age at transplantation was not associated with increased graft loss by 1-yr after transplantation. This is in contrast to the intermediate (3-yrs) and especially long-term (10-yrs) after transplantation where there was a marked increase in graft loss in age groups with higher IGF-1 levels (adolescence and young adulthood). Importantly, the association of IGF-1 level with age-associated graft loss was present through all age groups from 0 to 60-yrs, with lower levels of graft loss when IGF-1 levels were lower (early in life and, especially, late in life) and *vice versa*. The fact that the association was not restricted to the adolescence age range suggests that the overall result is not related to medication non-compliance. The relative contributions of underlying biology and non-compliance will need to be carefully tested in a pediatric cohort that is outside the scope of this report.

One clinical question arising from this work is why the single remaining kidney in the living kidney donor does not experience a higher kidney failure risk due to the GH-IGF-1 axis effect. This apparent protection might arise from the fact that the donor has a relatively high 50% kidney dose remaining, which is predicted to impose a relatively small risk by 10yrs of follow-up (Figure 4 top panels). Other potential mitigating factors include the absence of an alloimmune milieu, lower GH levels in live donors (versus CKD and ESKD which are a high GH states)(51–53), and the presence of innervation that may protect against hyperfiltration. Regardless, current data suggests that there is indeed a relative increase in donor risk of ESKD in live kidney donors observable by 15-years after nephrectomy although this absolute risk is quite small(85). However concern remains that the risk of CKD and ESKD will increase as longer-term data is accumulated(86). These observations point to the need for careful donor follow-up and mitigation of compounding factors such as hypertension, obesity, and diabetes and possibly also IGF-1 level that amplify the risk for nephron loss(87).

One prediction from our clinical cohort data is that increasing the transplanted kidney dose will minimize the impact of high IGF-1 levels on IGF-1 exposure and thus improve allograft outcomes (e.g., particularly during adolescence and in younger adults) or other allograft recipients with high IGF-1, as suggested by our patient-level data. Conversely, in older recipients who tend to have lower IGF-1 levels or other individuals with low IGF-1 levels, a smaller kidney dose, including kidneys with some level of glomerulosclerosis that are currently discarded(88,89), might be expected to have good long-term survival, thereby increasing the pool of available kidneys, a top 3 goal for Advancing American Kidney Health(90). A second prediction is that reducing early allograft exposure to GH/IGF-1 will improve long-term allograft survival. ACE inhibition, by reducing hyperfiltration and thereby limiting graft exposure to GH/IGF-1, would be expected to be protective. Several ACE inhibitor trials in KTx have shown little clinical benefit(91–93). However, recent evidence suggests that very early versus later drug initiation may improve tissue injury and long-term function surrogates(92,94). Our animal model and clinical data raises the possibility that the lack of an observed ACE inhibition protective effect could occur because the drug was not started until well after transplantation due to concern for potential side effects (hyperkalemia, reduced renal perfusion, hypotension, and anemia)(95). Our modeling data suggests that the significant growth events that ultimately determine allograft destiny occur very early after transplantation, and thus, initiating ACEi outside of this window minimizes the potential benefit of early ACEi(16,27). We speculate that alternative approaches to extending allograft survival include specific IGF-1R blockade (currently in use to treat immune-mediated thyroid eye disease) and/or targeting GH (by GHRH inhibition using octreotide or its analogs) may be feasible(96). However, long-term use of these agents may be limited, given their side effects and critical functions of GH/IGF-1, especially during rapid body growth. Thus, interventions to modulate circulating IGF-1 levels and/or reduce allograft exposure to IGF-1 signaling may enhance allograft longevity.

While we provide substantial evidence for the role of IGF-1 signaling in KTx, unequivocal proof that the GH-IGF-1 axis drives shorter graft survival will require prospective clinical trials that specifically limit allograft exposure to IGF-1. Another limitation of this report is that kidney dose was extrapolated through the donor-to-recipient BSA ratio, which also serves as a surrogate for its projected hyperfiltration effect(97). However, the use of this surrogate is supported by the strong correlation between body surface area (BSA) and kidney size(98,99). A further limitation of the transcriptomic data is that the time 0 and biopsies in the first-year allografts were not paired. However, reassuringly, the findings in the transcriptomic analysis are consistent with findings in previous Uni-Nx model systems(28,50).

Taken together we highlight a novel association between circulating IGF-1 levels at the time of KTx and long-term allograft survival. Figure 9 summarizes putative mechanisms for how kidney GH-IGF-1 pathways become dysregulated in allografts, leading to diverse pathologic processes that together shorten kidney lifespan.

**Figure 9:**
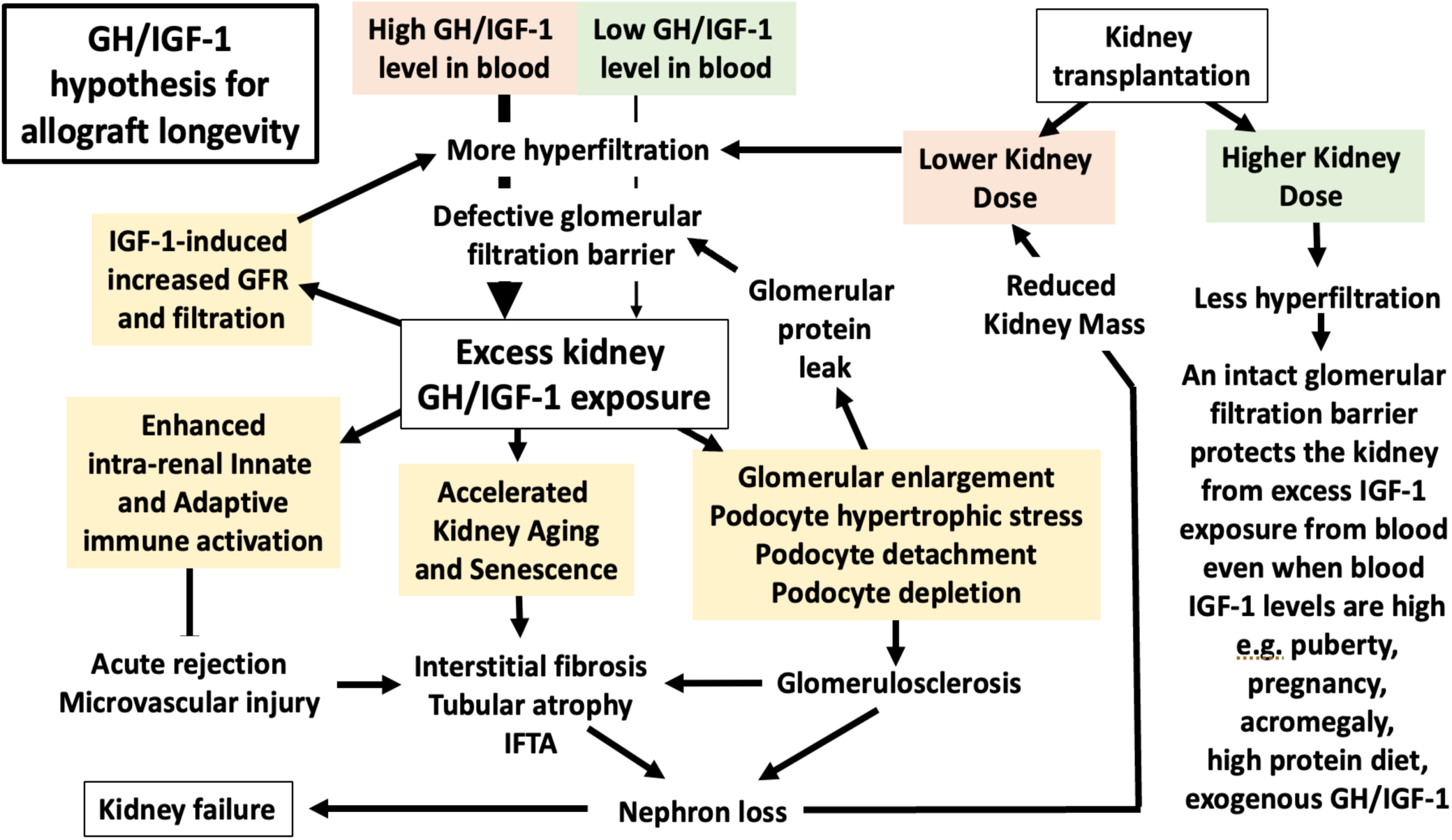
The single kidney state sets the allograft up for increased IGF-1 exposure dependent on the donor kidney dose and, thereby, the extent of hyperfiltration. A higher IGF-1 exposure, with its potential diverse downstream consequences, is shown in yellow boxes. We hypothesize that in addition to its role in early glomerular hypertrophy and podocyte stress(11–13,16), IGF-1 is likely to amplify the glomerular filtration rate by a nitric-oxide mediated vasodilatory response(3) with higher IGF-1 levels leading to a higher vasodilatory response. These data are also supported by the observations that an IGF-1R and nitric oxide inhibitor(3,108,109) can mitigate hyperfiltration. Furthermore, IGF-1 could promote fibroblast activation in KTx as it does in other model systems(110,111), leading to the development of Interstitial Fibrosis and Tubular Atrophy (IFTA) purported to be a final common pathway of graft loss(112). In addition, IGF-1 is known to enhance immune activation, which was also supported in our analysis, where an increased risk of a T-cell T-cell-mediated rejection was observed. The Insulin-Insulin-like growth factor system (IIS) drives shorter lifespans in worms, flies, and mice. We speculate that similar IGF-1 signaling via IGF-1R may also accelerate senescence, which is reported in kidney allografts. As noted by our analysis, larger kidney doses reduce kidney exposure to hyperfiltered IGF-1, thereby relatively protecting the normal two kidney state from the downstream effects of high circulating IGF-1 levels under different conditions (e.g., during puberty and diseases associated with high IGF-1 levels, including acromegaly, pregnancy, and exogenous GH and IGF-1 treatments).

## Methods

### Cohort 1: Population-Level Study

Data from the Organ Procurement and Transplantation Network (OPTN) database (October 1, 1987, to March 31, 2022) was analyzed. After applying exclusion criteria (**Supplementary Figure S1**), 366,404 observations remained for analysis. Incident DCGF rate for recipients ages 0-60 years was determined using the Kaplan-Meier estimator, censoring for death, re-transplantation, loss to follow-up, or the end of the follow-up period. The "stptime" command was used to generate incident DCGF rates per 1000 patient-years by age at KTx and also age at time of DCGF. he assumption of Cox proportional hazards was verified using Schoenfeld residuals, with the Stata command ‘estat phtest’ employed for this purpose. The incident DCGF rate was also parsed by time post-KTx to generate incident DCGF rates for 1-year, 3-year, and 10-year outcomes. Furthermore, we obtained the time by which 50% of allografts are lost, censoring for death for each age group to obtain a "median allograft survival" by age. The IGF-1 level for each age group for males and females was obtained from population-level average serum IGF-1 levels by age and gender across 0-60 years, obtained from Bidlingmaier et al. (n=15,014). A Welch’s t-test assuming unequal variances for males and females was used to assess the overall difference in mean IGF-1 levels between males and females and was not statistically significant.

### Cohort 2: Clinical Study

All human subject work was carried out per the code of ethics of the world medical association (Declaration of Helsinki) and conformed with the Declaration of Istanbul. This study received approval from the Institutional Review Board at the University of Michigan (IRB# HUM00228292). Serum samples stored before KTx were analyzed for 216 consecutive recipients who underwent KTx at this institution starting July 1, 2013. Luminex Assay was utilized to measure total IGF-1 and IGF-2 levels using MILLIPLEX MAP human IGF-I and IGF-II magnetic bead panels (EMD Millipore, Billerica, MA) in duplicate.

#### Power analysis

A sample size estimation of 211 patients was calculated using a hazard ratio of 2.0, an event probability of 0.31, and a power of 0.8. The event probability was based on an interim analysis of the initial 106 recipients. The hazard ratio of 2.0 used for analysis is a conservative effect size compared to the hazard ratios from previously reported uni-nephrectomy animal data, which ranged from 4-8-fold.(4,5,100)

##### Effective Kidney Dose (eKD)

We calculated an "Effective Kidney Dose" (eKD) for each recipient using the donor-to-recipient body surface area (BSA) ratio to modify the value of 50% of the normal kidney dose that is routinely provided by a single kidney transplant that replaces the function normally provided by two kidneys. The donor-to-recipient BSA ratio has long been used in KTx studies to estimate the donor kidney-to-recipient size mismatch studies(101,102). For ease of interpretation and analysis, the ratio was converted into a percentage, representing the ideal two-kidney complement for that recipient’s body size. For example, if a donor and recipient have the same BSA, the ratio would be 1, which equates to 50% of the ideal two-kidney complement. A Donor BSA > Recipient BSA means that >50% eKD has been delivered (greater than 1-kidney complement), and if the Donor BSA < Recipient BSA, then the eKD delivered will be <50% (less than 1-kidney complement).

##### Allograft IGF-1 exposure

Allograft IGF-1 exposure is a function of both the IGF-1 concentration in the recipient’s blood at time of transplantation and the effective kidney dose (eKD) delivered, which has long been used as a surrogate for the amount of hyperfiltration(8). Thus, we modeled allograft IGF-1 exposure at KTx by the IGF-1 level and its interaction with the kidney dose (IGF-1*eKD). Figure 2 provides a visual representation of how Allograft IGF-1 exposure was modeled. Allograft IGF-2 exposure was similarly modeled.

##### Outcomes of Interest

The *primary endpoint* of this study was the time-to-death-censored Graft Failure (DCGF) from any cause. *Secondary endpoint*s included the development of proteinuria (defined as ≥1g/g of urinary creatinine) and the occurrence of Biopsy-Proven Acute Rejection (BPAR). *Sensitivity analyses* were performed to determine whether the effects of IGF-1 exposure on DCGF were independent of post-transplant complications such as BPAR(62), the emergence of *de novo* donor-specific antibodies (*dnDSA*), or predictors of pre-transplant allo-sensitization including the number of previous transplants and calculated panel reactive antibodies (cPRA) for HLA Class I or II. *Additional sensitivity analyses* were performed on the BPAR subgroup, specifically T-cell mediated rejection (TCMR)(62). Due to the low number of events in the antibody-mediated rejection and mixed rejection categories, no additional sensitivity analysis was performed.

##### Survival Analysis

We utilized multivariable Cox regression models to analyze the effect of IGF-1 exposure (IGF-1*eKD interaction) on primary and secondary study endpoints. These models were adjusted for known covariates influencing long-term outcomes, which were determined *a priori*.

##### Statistical Analysis

Continuous variables were presented as means ± standard deviations, and categorical variables were reported as proportions. The assumption of Cox proportional hazards was verified using Schoenfeld residuals, with the Stata command ‘estat phtest’ employed for this purpose. Post-multivariable Cox regression, the ‘stcurvè command was utilized to visualize covariate-adjusted outcomes across different IGF-1 values (90th, 50th, and 10th percentiles distribution) and eKD values (at the 1st, 10th, 25th, 50th, 75th, 90th, and 99th percentiles). The eKD was mean-centered to allow the interpretation of the independent IGF-1 effect on the outcomes of interest at the mean eKD in this cohort. All analyses used Stata/MP 17.0 (College Station, Texas) and GraphPad Prism

### Cohort 3: Genotype studies

Genotype-to-phenotype associations were explored through a meta-analysis of three published genotype/phenotype studies focused on KTx outcomes: GoCAR, Genomics in Chronic Allograft Rejection (GoCAR), and the Clinical Trials in Organ Transplantation (CTOT) 01 and 17(45,103,104). Multivariable Cox models, adjusted for clinical covariates, were developed in additive formats across the three cohorts. Due to the unavailability of donor weight data or body surface area (BSA), the models included recipient weight as a variable to account for the recipient’s body size.

### Cohort 4: Transcriptomic Analysis of IGF-1 Signaling in Healthy and Transplanted Kidneys

Dataset: Reference healthy human transcriptomic data from the Human Kidney and Transplant Transcriptome Atlas (HKTTA) were utilized to define the transcriptional landscape of the IGF-1 signaling pathway in a healthy binephric state(30,47).

Single-cell processing of the research core: The study uses data previously published by Menon et al. (Menon et al., 2022)(30). The generation of single-cell preparations was accomplished by enzymatic (Liberase TL; Sigma Aldrich) and mechanical dissociation for 12 minutes at 37 °C following our previously published protocol(105). Cells are filtered through a 30 μm strainer and counted, and up to 10,000 viable cells are submitted to the University of Michigan Advanced Genomics Core facility to execute droplet-based high-throughput scRNA-seq on the 10x Genomics Chromium platform. After droplet encapsulation, the 10x Genomics approach allows cell lysis, individual cell RNA molecular barcoding, and reverse transcription. Subsequently, cDNA libraries are generated and sequenced on the NovaSeq 6000 platform (Illumina) as asymmetric paired-end (26 × 151) runs and generating >200 million raw sequence reads per sample. The sequencing data are preprocessed using the 10x Genomics software Cell Ranger. Downstream analysis was performed with the Seurat R package version 3 (Satija Lab)(106) A combined analysis of the single-cell data sets generated from the different sample sources (living donor and surveillance biopsies) using Seurat version 3 includes the following steps: filtering of cells with <500 genes or >50% mitochondrial content default normalization, scaling based on sample mRNA count and mitochondrial RNA content, dimensionality reduction principal component analysis and uniform manifold approximation and projection, sample integration using the Harmony algorithm, standard unsupervised clustering, and the discovery of differentially expressed cell type-specific markers. Cell-type specific differential gene expression analyses (DEGs) were performed using a pseudobulk approach, which was employed to mitigate the risk of false positive discovery commonly associated with single-cell transcriptomic studies(107). The signaling elements tested, determined *a priori* from the literature, included transcripts for *IGF1, IGF2*, IGF-1 receptor (*IGF1R*), growth hormone receptor (*GHR*), and IGF binding proteins (*IGFBPs 1-6*)(73).

#### Pseudobulking strategies

Pseudobulking strategies were applied to minimize the risk of false positive results in single-cell data, especially among cells with low counts(107).After clustering and cell type identification, the pseudo-bulk matrices were generated from the single-cell matrix (25,720 genes as rows, 40,254 cells as columns) by aggregating cell-wise raw counts into their corresponding cell type/sample bins using summation. These data have been previously published in Menon et al. and are available on the Gene Expression Omnibus, Accession number GSE169285(30). From 34 samples and 29 cell types, this yielded a matrix with 965 cell type/sample columns, excluding 31 representing cases where cell types did not occur in a given sample. To facilitate cell-type-specific analysis over samples, 29 cell-type-specific matrices, each with genes as rows and samples as columns, were extracted along corresponding sample-related metadata. This process conserved the number of counts in the original single-cell data exactly, leaving all QC-related filtering of genes and samples to downstream analysis. Generally, as some cell types are much more abundant than others – due both to their varied frequency in the original biopsy samples and recovery biases in the single-cell protocol itself (e.g., from dissociation efficiencies) – and sequencing coverage per cell also varies widely, some samples in some cell types were of insufficient quality (sequencing depth) to support analysis and were excluded. For differential expression analysis, a sample was excluded in a particular cell type if it was supported by fewer than 10 cells or 10,000 total counts. At this point, each cell type-specific counts matrix could be treated similarly to one derived from bulk RNA-seq, with normalization done using limma voom and differential expression with limma.

## Data Availability

All data produced in the present study are available upon reasonable request to the authors

## Acknowledgements

**JAB:** JAB acknowledges support from the NIH (grants K08 DK125776).

**PH:** PH acknowledges support from the NIH (U01 AI 063594).

**MCM:** MCM acknowledges support from the NIH (grants R01DK122164, R01DK132274, and R21AI178705) and the Department of Defense Awards (Awarding Office: USAMRAA under HT94252310454, HT94252310441).

**MK:** MK is partially supported for this work through the George M. O’Brien Michigan Kidney National Resource Center, which is funded by NIH/NIDDK grant U54DK137314 and the previous grant, the University of Michigan O’Brien Kidney Translational Core Center P30 DK081943.

**RCW:** RCW acknowledges prior support from NIH grants R01 DK46073 and R01 DK102643 and the University of Michigan O’Brien Kidney Translational Core Center P30 DK081943.

**ASN:** ASN acknowledges support of Intramural grants from the University of Michigan O’Brien Kidney Translational Core Center P30 DK081943, First Pathway Award, Michigan Institute of Clinical Health Research UL1TR002240, Startup funds from the Department of Internal Medicine; Taubman Institute and an extramural grant from the National Institute of Health K23 DK 125529.

This work was partly supported by the Health Resources and Services Administration contract HHSH250-2019-00001C. The content is the responsibility of the authors alone and does not necessarily reflect the views or policies of the Department of Health and Human Services, nor does mention of trade names, commercial products, or organizations imply endorsement by the US Government. This work was supported in part by the Health Resources and Services Administration contract HHSH250-2019-00001C. The content is the responsibility of the authors alone and does not necessarily reflect the views or policies of the Department of Health and Human Services, nor does mention of trade names, commercial products, or organizations imply endorsement by the US Government.

The (NEPTUNE) is part of the Rare Diseases Clinical Research Network (RDCRN), which is funded by the National Institutes of Health (NIH) and led by the National Center for Advancing Translational Sciences (NCATS) through its Division of Rare Diseases Research Innovation (DRDRI). NEPTUNE is funded under grant number U54DK083912 as a collaboration between NCATS and the National Institute of Diabetes and Digestive and Kidney Diseases (NIDDK). Additional funding and/or programmatic support is provided by the University of Michigan, NephCure Kidney International, Alport Syndrome Foundation, and the Halpin Foundation. RDCRN consortia are supported by the RDCRN Data Management and Coordinating Center (DMCC), funded by NCATS and the National Institute of Neurological Disorders and Stroke (NINDS) under U2CTR002818.

## Author Contributions

RCW and ASN developed the concept jointly for >8 years, building on RCW’s podocyte depletion hypothesis as it applied to the single kidney state and kidney transplantation. ASN and RCW planned the studies. ASN and RCW jointly wrote the first and final drafts of the manuscript. MC identified and pulled samples for analysis, performed IGF-1 and IGF-2 assays, and provided critical feedback regarding sample QI and planning of the individual patient study. All data analysis was performed by ASN, VN, DF, and JH and interpreted by ASN, RCW, JAB, VN, PH, MCM, MK, and MC. CTOT and GoCAR data was kindly provided by PH, MCM, ZS, and ZZ; MCM. SN, ND, and KS collated all the clinical data. All authors agree to the final version of this manuscript and consented to submission.

## Data Statement

The OPTN data is publicly available at request from OPTN. All other de-identified clinical data can be obtained at request to.

## Supplementary Materials

### Methods

#### Transcriptomics

Single-cell processing of the research core.

Psueudobulking strategies

### Figures

Supplementary Figure 1: Patient-level data from the Individual Patient Study.

Supplementary Figure 2: Flowchart of how population-level study was analyzed

## Tables

**Supplementary Table 1:** Relationship between IGF-1 exposure (IGF-1*eKD interaction) and Death Censored Graft Failure.

| **Variables** | **Haz. ratio** | **Std. err.** | **P value** | **LCL** | **UCL** |
| --- | --- | --- | --- | --- | --- |
| IGF-1 | 0.844 | 0.087 | 0.099 | 0.690 | 1.033 |
| eKD | 1.014 | 0.015 | 0.334 | 0.986 | 1.043 |
| IGF-1*eKD | 0.985 | 0.005 | 0.007 | 0.974 | 0.996 |
| Recipient Age | 0.963 | 0.013 | 0.006 | 0.938 | 0.989 |
| Recipient Gender (Male) | 1.106 | 0.392 | 0.776 | 0.552 | 2.216 |
| Recipient Race (vs White) |  |  |  |  |  |
| African American | 3.156 | 1.153 | 0.002 | 1.542 | 6.458 |
| Others | 1.902 | 1.430 | 0.392 | 0.436 | 8.300 |
| Donor Age | 1.043 | 0.013 | 0.001 | 1.019 | 1.069 |
| Donor Race (vs. White) |  |  |  |  |  |
| African American | 2.845 | 1.454 | 0.041 | 1.044 | 7.749 |
| Others | 3.279 | 1.746 | 0.026 | 1.155 | 9.310 |
| Donor Gender (Male) | 1.184 | 0.405 | 0.622 | 0.605 | 2.316 |
| Decreased Donor | 3.511 | 2.184 | 0.043 | 1.037 | 11.880 |
| Cold Ischemia time (min) | 1.000 | 0.001 | 0.992 | 0.999 | 1.001 |

**IGF-1:** Insulin-like Growth Factor 1; **eKD:** Estimated Kidney Dose

**Supplementary Table 2:** Relationship between IGF-1 exposure (IGF-1*eKD interaction) and Death Censored Graft Failure also adjusted for pretransplant sensitization using variables Cpra and previous transplantation.

| **Variables** | **Haz. ratio** | **Std. err.** | **P value** | **LCL** | **UCL** |
| --- | --- | --- | --- | --- | --- |
| IGF-1 | 0.840 | 0.085 | 0.082 | 0.689 | 1.023 |
| eKD | 1.017 | 0.014 | 0.238 | 0.989 | 1.045 |
| IGF-1*eKD | 0.985 | 0.005 | 0.007 | 0.975 | 0.996 |
| Recipient Age | 0.965 | 0.013 | 0.007 | 0.940 | 0.990 |
| Recipient Gender (Male) | 0.911 | 0.346 | 0.806 | 0.433 | 1.917 |
| Recipient Race (vs White) |  |  |  |  |  |
| African American | 3.153 | 1.191 | 0.002 | 1.504 | 6.610 |
| Others | 1.485 | 1.131 | 0.603 | 0.334 | 6.608 |
| Donor Age | 1.046 | 0.014 | 0.001 | 1.019 | 1.073 |
| Donor Race (vs. White) |  |  |  |  |  |
| African American | 3.532 | 1.882 | 0.018 | 1.243 | 10.038 |
| Others | 4.303 | 2.406 | 0.009 | 1.439 | 12.873 |
| Donor Gender (Male) | 1.100 | 0.381 | 0.784 | 0.558 | 2.168 |
| Decreased Donor | 3.676 | 2.414 | 0.047 | 1.015 | 13.313 |
| Cold Ischemia time (min) | 1.000 | 0.001 | 0.798 | 0.999 | 1.001 |
| Cpra Class 1 | 0.983 | 0.009 | 0.075 | 0.965 | 1.002 |
| Cpra Class 2 | 1.004 | 0.010 | 0.732 | 0.983 | 1.024 |
| Previous TP | 0.658 | 0.387 | 0.477 | 0.208 | 2.086 |

**IGF-1:** Insulin-like Growth Factor 1; **eKD:** Estimated Kidney Dose; **TP:** Transplantation; **Cpra:** Calculated Panel Reactive Antibodies

**Supplementary Table 3:** Relationship between IGF-1 exposure (IGF-1*eKD interaction) and Death Censored Graft Failure also adjusted for the presence of post-transplant *de novo* donor-specific antibody development (either Class 1, Class 2, or both)

| Variables | Haz. ratio | Std. err. | P value | LCL | UCL |
| --- | --- | --- | --- | --- | --- |
| IGF-1 | 0.833 | 0.085 | 0.075 | 0.682 | 1.018 |
| eKD | 1.010 | 0.014 | 0.478 | 0.982 | 1.039 |
| IGF-1*eKD | 0.986 | 0.005 | 0.008 | 0.975 | 0.996 |
| Recipient Age | 0.960 | 0.014 | 0.004 | 0.934 | 0.987 |
| Recipient Gender (Male) | 1.155 | 0.417 | 0.691 | 0.569 | 2.343 |
| Recipient Race (vs White) |  |  |  |  |  |
| African American | 2.958 | 1.116 | 0.004 | 1.412 | 6.195 |
| Others | 2.189 | 1.697 | 0.312 | 0.479 | 10.001 |
| Donor Age | 1.044 | 0.013 | 0.000 | 1.019 | 1.069 |
| Donor Race (vs. White) |  |  |  |  |  |
| African American | 2.769 | 1.404 | 0.045 | 1.024 | 7.482 |
| Others | 3.038 | 1.637 | 0.039 | 1.057 | 8.734 |
| Donor Gender (Male) | 1.149 | 0.400 | 0.691 | 0.580 | 2.274 |
| Decreased Donor | 3.624 | 2.297 | 0.042 | 1.046 | 12.552 |
| Cold Ischemia time (min) | 1.000 | 0.001 | 0.983 | 0.999 | 1.001 |
| Donor Specific Antibody | 1.439 | 0.509 | 0.303 | 0.719 | 2.880 |

**IGF-1:** Insulin-like Growth Factor 1; **eKD:** Estimated Kidney Dose

**Supplementary Table 4:** Relationship between IGF-1 exposure (IGF-1*eKD interaction) and Death Censored Graft Failure adjusted for post-transplant Biopsy Proven Acute Rejection.

| **Variables** | **Haz.**<br>**ratio** | **Std. err.** | **P value** | **LCL** | **UCL** |
| --- | --- | --- | --- | --- | --- |
| IGF-1 | 0.866 | 0.092 | 0.177 | 0.702 | 1.067 |
| eKD | 1.008 | 0.014 | 0.544 | 0.982 | 1.035 |
| IGF-1*eKD | 0.986 | 0.005 | 0.009 | 0.975 | 0.996 |
| Recipient Age | 0.965 | 0.014 | 0.012 | 0.938 | 0.992 |
| Recipient Gender (Male) | 1.039 | 0.382 | 0.918 | 0.506 | 2.134 |
| Recipient Race (vs White) |  |  |  |  |  |
| African American | 4.008 | 1.742 | 0.001 | 1.710 | 9.395 |
| Others | 2.937 | 2.336 | 0.175 | 0.618 | 13.960 |
| Donor Age | 1.064 | 0.016 | 0.000 | 1.034 | 1.095 |
| Donor Race (vs. White) |  |  |  |  |  |
| African American | 2.075 | 1.125 | 0.178 | 0.717 | 6.003 |
| Others | 1.322 | 0.826 | 0.655 | 0.388 | 4.500 |
| Donor Gender (Male) | 1.023 | 0.385 | 0.951 | 0.490 | 2.138 |
| Decreased Donor | 3.284 | 2.222 | 0.079 | 0.872 | 12.372 |
| Cold Ischemia time (min) | 1.000 | 0.001 | 0.943 | 0.999 | 1.001 |
| Type of Rejection |  |  |  |  |  |
| TCMR | 5.018 | 2.458 | 0.001 | 1.921 | 13.105 |
| AMR | 6.597 | 4.216 | 0.003 | 1.885 | 23.085 |
| Mixed | 4.201 | 2.857 | 0.035 | 1.107 | 15.934 |

**IGF-1:** Insulin-like Growth Factor 1; **eKD:** Estimated Kidney Dose; **TCMR:** T-Cell Mediated Rejection; **AMR:** Antibody Mediated Rejection

**Supplementary Table 5:** Relationship between IGF-1 exposure (IGF-1*eKD interaction) and proteinuria of >1g/g on follow-up.

| **Variables** | **Haz. ratio** | **Std. err.** | **P value** | **LCL** | **UCL** |
| --- | --- | --- | --- | --- | --- |
| IGF-1 | 0.952 | 0.091 | 0.607 | 0.789 | 1.149 |
| eKD | 1.021 | 0.016 | 0.186 | 0.990 | 1.053 |
| IGF-1*eKD | 0.987 | 0.005 | 0.013 | 0.976 | 0.997 |
| Recipient Age | 0.972 | 0.014 | 0.055 | 0.944 | 1.001 |
| Recipient Gender (Male) | 0.934 | 0.383 | 0.868 | 0.418 | 2.086 |
| Recipient Race (vs White) |  |  |  |  |  |
| African American | 2.147 | 0.906 | 0.070 | 0.940 | 4.908 |
| Others | 1.082 | 0.990 | 0.932 | 0.180 | 6.508 |
| Donor Age | 1.025 | 0.014 | 0.072 | 0.998 | 1.054 |
| Donor Race (vs. White) |  |  |  |  |  |
| African American | 3.929 | 2.241 | 0.016 | 1.285 | 12.015 |
| Others | 2.826 | 1.708 | 0.086 | 0.864 | 9.241 |
| Donor Gender (Male) | 1.384 | 0.526 | 0.392 | 0.657 | 2.915 |
| Decreased Donor | 1.390 | 0.971 | 0.638 | 0.353 | 5.465 |
| Cold Ischemia time (min) | 1.001 | 0.001 | 0.446 | 0.999 | 1.002 |

**Supplementary Table 6:** Relationship between IGF-1 exposure (IGF-1*eKD interaction) and Biopsy Proven Acute Rejection.

| Variables | Haz. ratio | Std. err. | P value | LCL | UCL |
| --- | --- | --- | --- | --- | --- |
| IGF-1 | 0.881 | 0.074 | 0.133 | 0.748 | 1.039 |
| eKD | 1.006 | 0.012 | 0.599 | 0.983 | 1.030 |
| IGF-1*eKD | 0.994 | 0.003 | 0.068 | 0.987 | 1.000 |
| Recipient Age | 0.968 | 0.014 | 0.019 | 0.941 | 0.995 |
| Recipient Gender (Male) | 0.820 | 0.316 | 0.606 | 0.385 | 1.744 |
| Recipient Race (vs White) |  |  |  |  |  |
| African American | 0.792 | 0.315 | 0.557 | 0.363 | 1.727 |
| Others | 0.744 | 0.496 | 0.658 | 0.202 | 2.749 |
| Donor Age | 0.974 | 0.013 | 0.047 | 0.949 | 1.000 |
| Donor Race (vs. White) |  |  |  |  |  |
| African American | 6.105 | 3.104 | 0.000 | 2.253 | 16.539 |
| Others | 7.388 | 3.328 | 0.000 | 3.056 | 17.863 |
| Donor Gender (Male) | 1.216 | 0.433 | 0.583 | 0.605 | 2.443 |
| Decreased Donor | 1.963 | 1.167 | 0.257 | 0.612 | 6.295 |
| Cold Ischemia time (min) | 1.000 | 0.001 | 0.824 | 0.999 | 1.001 |
| Cpra 1 | 1.001 | 0.009 | 0.916 | 0.984 | 1.018 |
| Cpra 2 | 1.005 | 0.010 | 0.610 | 0.986 | 1.025 |
| Previous TP | 0.916 | 0.442 | 0.855 | 0.356 | 2.356 |

**IGF-1:** Insulin-like Growth Factor 1; **eKD:** Estimated Kidney Dose; **TP:** Transplantation; **Cpra:** Calculated Panel Reactive Antibodies

**Supplementary Table 7:** Relationship between IGF-1 exposure (IGF-1*eKD interaction) and T-Cell Mediated Rejection.

| **Variables** | **Haz.**<br>**ratio** | **Std. err.** | **P value** | **LCL** | **UCL** |
| --- | --- | --- | --- | --- | --- |
| IGF-1 | 0.787 | 0.098 | 0.055 | 0.616 | 1.005 |
| eKD | 1.005 | 0.020 | 0.791 | 0.968 | 1.044 |
| IGF-1*eKD | 0.985 | 0.007 | 0.044 | 0.970 | 1.000 |
| Recipient Age | 0.957 | 0.018 | 0.016 | 0.923 | 0.992 |
| Recipient Gender (Male) | 0.371 | 0.180 | 0.041 | 0.144 | 0.960 |
| Recipient Race (vs White) |  |  |  |  |  |
| African American | 0.460 | 0.271 | 0.187 | 0.145 | 1.459 |
| Others | 1.924 | 1.481 | 0.395 | 0.426 | 8.695 |
| Donor Age | 0.980 | 0.016 | 0.219 | 0.950 | 1.012 |
| Donor Race (vs. White) |  |  |  |  |  |
| African American | 11.617 | 8.401 | 0.001 | 2.815 | 47.932 |
| Others | 10.051 | 5.117 | 0.000 | 3.706 | 27.260 |
| Donor Gender (Male) | 1.391 | 0.621 | 0.459 | 0.580 | 3.335 |
| Decreased Donor | 3.081 | 2.380 | 0.145 | 0.678 | 14.006 |
| Cold Ischemia time (min) | 1.000 | 0.001 | 0.624 | 0.998 | 1.001 |
| Cpra 1 | 0.983 | 0.012 | 0.180 | 0.959 | 1.008 |
| Cpra 2 | 1.015 | 0.013 | 0.225 | 0.991 | 1.040 |
| Previous TP | 1.374 | 0.672 | 0.516 | 0.527 | 3.583 |

**IGF-1:** Insulin-like Growth Factor 1; **eKD:** Estimated Kidney Dose; **TP:** Transplantation; **Cpra:** Calculated Panel Reactive Antibodies

**Supplementary Table 8:** Genotype of patients with the SNP rs35767 categorized by self-reported race in the NEPTUNE Study among 620 NEPTUNE patients.

| **Self-Reported Race** | **REF** | **HET** | **HOM_ALT** | **Total** | **NEPTUNE AF** |
| --- | --- | --- | --- | --- | --- |
| Asian/Asian American | 9 | 23 | 30 | 62 | 0.6694 |
| Black/African American | 19 | 85 | 43 | 147 | 0.5816 |
| Multi-Racial | 2 | 15 | 15 | 32 | 0.7031 |
| Native American/Alaskan<br>Native/First Nation | 0 | 1 | 0 | 1 | 0.5 |
| Native Hawaiian/Other Pacific<br>Islander | 0 | 2 | 2 | 4 | 0.75 |
| Unknown | 4 | 21 | 21 | 46 | 0.6848 |
| White/Caucasian | 9 | 86 | 233 | 328 | 0.8415 |

**Supplement Figure 1:**
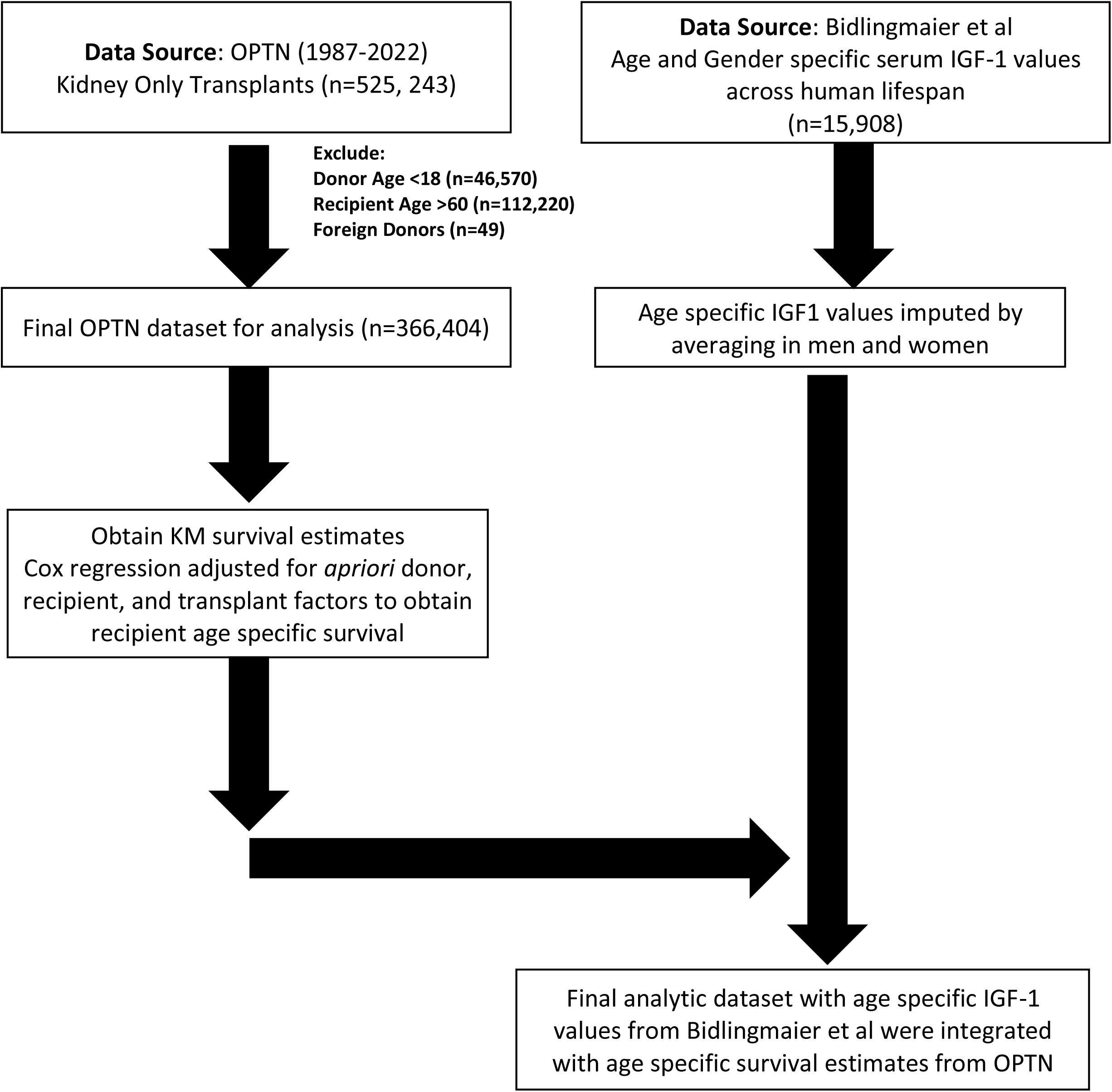
Integration of OPTN Kidney Transplant Data with Age-Specific IGF-1 Levels This figure illustrates the methodology and data integration process used in our study. We analyzed the Organ Procurement and Transplantation Network (OPTN) dataset spanning from 1987 to 2022, focusing exclusively on patients who received kidney-only transplants. Initial data included 525,243 patients; after excluding donors younger than 18, recipients older than 60, and foreign donors, the cohort was narrowed down to 366,404 patients. Concurrently, age and gender-specific serum IGF-1 levels across the human lifespan were obtained from the study by Bidlingmaier et al.^34^ Average age-specific IGF-1 values were calculated by averaging data for both genders. These values were then integrated with age-specific survival estimates derived from the OPTN dataset to assess the impact of IGF-1 levels on post-transplant outcomes. This figure provides a schematic representation of the data extraction and integration processes employed to facilitate this comprehensive analysis.

